# Deep Brain Stimulation of the Fornix for Alzheimer’s Disease: A Systematic Review and Meta-Analysis of Cognitive and Motor Outcomes

**DOI:** 10.64898/2025.11.30.25341330

**Authors:** Farzan Fahim, MohammadAmin Farajzadeh, Behnaz Rahatijafarabad, Abdul Majeed Mohammadi, Ali Khorram, Mehrsa Mostafaei, Omidreza Nikbakht, Raziyeh Zamiri, Nikoo Sepehrian, Sayeh Oveisi, Alireza Zali

**Author notes:** Corresponding author: Farzan Fahim. These authors contributed equally as first co-authors (Farzan Fahim, MohammadAmin Farajzadeh).

## Abstract

**Background:** Deep brain stimulation of the fornix (DBS-f) has emerged as a potential strategy for Alzheimer’s disease (AD), based on evidence that stimulating the Papez circuit may enhance hippocampal metabolism, increase hippocampal volume, and modulate large-scale memory networks. However, studies vary substantially in design, stimulation parameters, sample size, and disease stage, and the consistency of cognitive and motor effects remains unclear.

**Objective:** To systematically review and meta-analyze the cognitive outcome associated with fornix-targeted DBS in Alzheimer’s disease.

**Methods:** This systematic review followed PRISMA-2020 guidelines, and the protocol was registered in PROSPERO (ID: CRD420251175724). A comprehensive search was conducted in PubMed, Scopus, Embase, Web of Science (WOS) and Cochrane Database of Systematic reviews (CDSR) included randomized controlled trials (RCTs), cohort studies,and case series. Ten eligible studies were analyzed, including mild, moderate, and severe AD cohorts. Meta-analyses were performed for ADAS-Cog and MMSE outcomes using random-effects models. Subgroup analysis (observational vs RCT), trim-and-fill publication bias, sensitivity analysis, and meta-regression (age 63–65 years) were conducted. Structural and metabolic data (hippocampal volumetry and FDG-PET) and motor outcomes (FIM, ADL, Barthel Index) were narratively synthesized.

**Results:** Across included studies, cognitive outcomes showed highly variable short-term responses but no sustained improvement in controlled settings. Severe-AD cohorts demonstrated early gains, MMSE and MoCA improved in 1.5–3 months (Mao 2018), and dual-target fornix + NBM DBS yielded significantly higher MMSE at 3 months (p=0.002) and MoCA at 3 (p=0.003) and 12 months (p=0.010) (Xu 2024). In contrast, mild-AD RCTs showed no clinically meaningful benefit.

Meta-analysis demonstrated a null pooled effect for cognition: ADAS-Cog: SMD = 0.05 (95% CI spanning null), robust to trim-and-fill (k =1). MMSE: pooled SMD near zero, stable on leave-one-out sensitivity analysis. Subgroup comparisons (RCT vs observational) showed no differences (χ²=0.04–0.05, p>0.80), and meta-regression revealed no association between effect size and age (β = −0.04, p=0.33; τ²=0), confirming minimal between-study variance. Structural and metabolic findings consistently showed biological activation despite weak clinical response. Two patients demonstrated bilateral hippocampal volume increase at 12 months, and FDG-PET studies reported widespread metabolic increases across frontal–temporal–parietal–striatal–thalamic and frontal–temporal–parietal–occipital–hippocampal networks. Higher baseline metabolism and increased metabolism at one year correlated with less cognitive decline. Motor outcomes showed no sustained improvements. FIM scores improved significantly more in the DBS group at 3 months (p<0.05) but not at 12 months (p=0.968). ADL and Barthel scores showed mixed responses in small severe-AD samples. DBS parameters were heterogeneous (1–7 V; 60–210 μs; 130 Hz in most studies), and programming duration varied markedly, underscoring a lack of standardized neuromodulatory protocol.

**Conclusion:** Fornix DBS reliably activates limbic and memory-related circuits at a physiological level but does not provide consistent or sustained cognitive or motor benefits at currently used parameters. Evidence suggests age- and severity-dependent effects, with older or more advanced patients showing transient improvements, while mild AD patients do not benefit. Future research should prioritize precision targeting, biomarker-driven patient selection, optimized stimulation paradigms, and multi-target neuromodulation approaches.

## Introduction

Alzheimer’s disease (AD) is a major neurodegenerative disorder characterized by progressive cognitive decline, memory loss, and loss of functional independence. Its growing prevalence poses an enormous public health challenge worldwide. According to the World Alzheimer Report 2024, more than 55 million people globally are living with dementia, the majority with Alzheimer’s disease, projections suggest that this number could reach 150 million by 2050 due to population aging^1,2^. In addition to the cognitive deficits that the AD cause, The AD patients usually suffer from he motor and gait related disorders during the advancement of the disease, for example the balance impairments and muscle weakness is often during he late stages of the disease.

Despite decades of research, effective disease-modifying therapies remain limited. Currently available pharmacological treatments, such as cholinesterase inhibitors (e.g., donepezil, rivastigmine) and NMDA receptor antagonists (e.g., memantine), offer only modest symptomatic benefits and do not provide a treatment for the underlying pathophysiology of the disease^3^. Although anti-amyloid monoclonal antibodies have shown encouraging results, their high cost, variable clinical benefit, and safety concerns restrict their broad clinical use ^2^. These challenges have encouraged exploration of alternative and innovative strategies such as deep brain stimulation (DBS). DBS is an emerging neuro-modulatory approach that may improve memory and cognitive performance in individuals with mild to moderate impairment. It targets and modulates specific brain regions—including the nucleus basalis of Meynert, the fornix, and the ventral capsule/ventral striatum—to influence neural circuits involved in cognition ^4,15–17^. Based on its established efficacy in movement disorders, DBS has gained attention as a possible intervention for cognitive decline, particularly through stimulation of the fornix, which may enhance memory and learning in early-stage Alzheimer’s disease ^1,15,18^.

The fornix is one of the impartant components of Papez circuit, and it is crucial for memory processing. Damage or atrophy of this pathway is strongly associated with memory deficits in AD. Both human and animal studies have shown that fornix DBS can enhance hippocampal metabolism^15,16,19^, reduce amyloid burden, and in some cases reverse hippocampal atrophy ^15,20^. These findings suggest that modulating fornix activity might influence disease progression and cognitive outcomes. And stimulating fornix can increase the neural activity in the other related areas to the memory-circuit and hypothetically can slow the development of the disease^16,21–23^.

Despite the increasing interest in fornix DBS, published studies vary substantially in design, sample size, outcome measures, and duration of follow-up. The evidence regarding its cognitive and motor effects remains limited^15,18, 22–24^. This systematic review therefore aims to summarize the current data on fornix-targeted DBS in individuals with Alzheimer’s disease, focusing on its therapeutic potential, methodological challenges, and future research directions.

## Method

This systematic review was performed based on Preferred Reporting Items for Systematic Reviews and Meta-Analysis (PRISMA) 2020 guidelines and its protocol was registered in the International Prospective Register of Systematic Reviews (ID: CRD420251175724)

### Search strategy

A comprehensive literature search was conducted in PubMed, Scopus, Embase, Web of Science (WOS) and Cochrane Database of Systematic reviews (CDSR) to identify all relevant studies, without any date or language restrictions. If any non-English studies were identified, their exact translation into English was planned. The search strategy was designed to capture studies which used DBS in fornix for Alzheimer disease patients and compare it with other treatment options including DBS in other areas or conservative treatments.

For PubMed, the following search terms were used:

(Alzheimer* OR “Alzheimer disease” OR dementia OR “cognitive decline” OR “mild cognitive impairment") AND ("deep brain stimulation” OR DBS OR “brain stimulation” OR “neuromodulation") AND (fornix OR “fornical region” OR “fornical area”) AND (cognit* OR memory OR “executive function” OR “neuropsychological test*” OR “neurocognitive” OR “cognitive outcome*” OR “cognitive function*”)

Equivalent search strategies were adapted for Scopus, Embase, and Web of Science using their respective indexing terms and syntax. The detailed database-specific strategies are provided in Supplementary 1.

In total, 351 studies were found: 101 from Embase, 85 from Web of Science, 92 from Scopus, and 73 from PubMed. 190 duplicates were identified manually.

Titles and abstracts were screened by (ON and MM), resulting in 40 articles selected for full-text review. full-text articles (n=40) were independently reviewed by two reviewer(AM,AK) for screening .in this step, 2 excluded sheets including: study ID, DOI, Region and Reason of exclusion and, also, 2 primary included sheets including study ID, DOI, Region and “P-I-C-O-S” as columns by each reviewer were designed, which attached as supplementary file 2. PRISMA flowchart summarizing the study selection process is provided in Figure 1.

**Figure 1.**
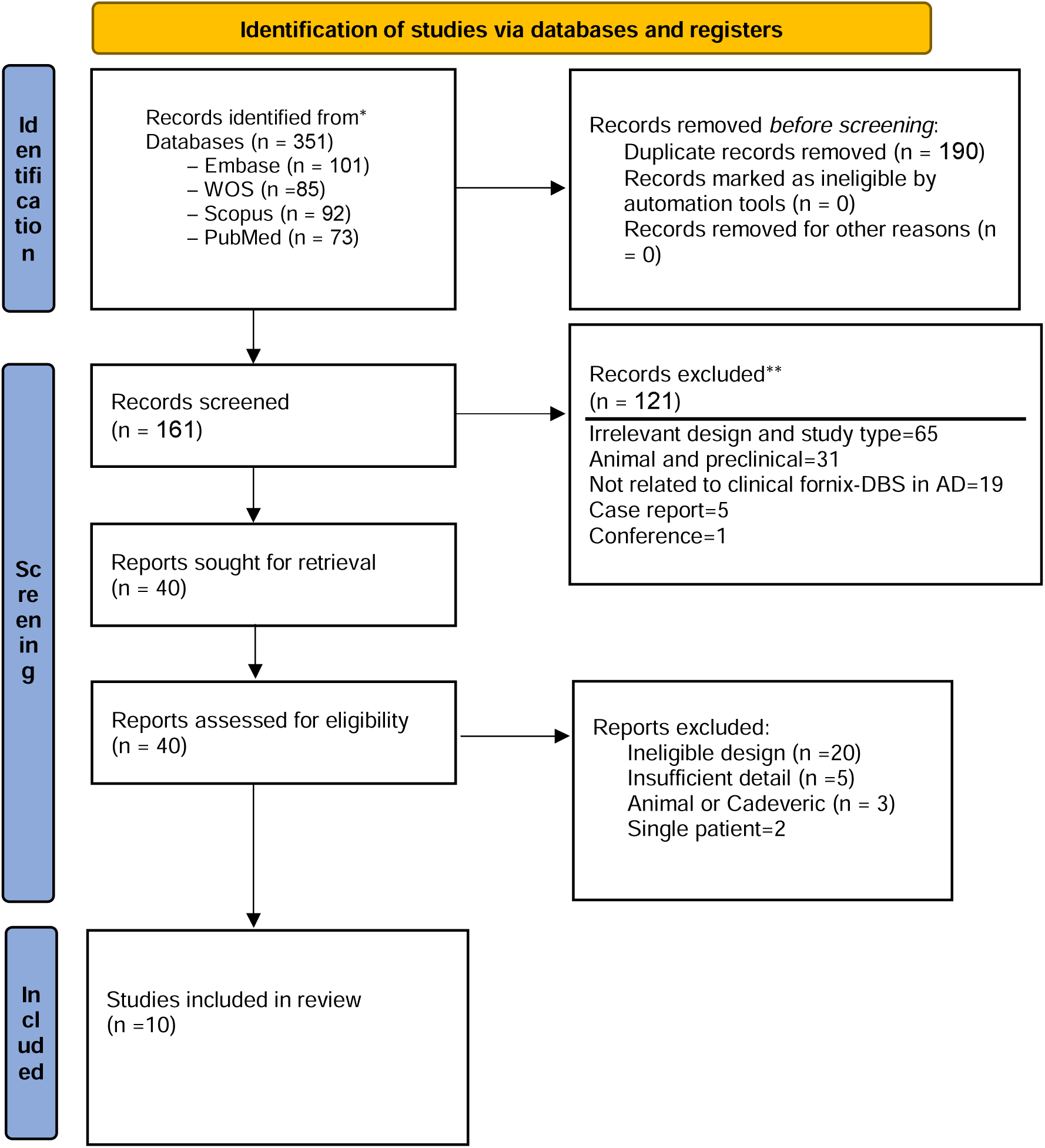
PRISMA flow diagram of study selection. A total of 351 records were identified across four databases and 190 duplicates were removed. After title and abstract screening, 40 articles underwent full-text review by two independent reviewers. And 10 studies included in the final stage. Source: Page MJ, et al. BMJ 2021;372:n71. doi: 10.1136/bmj.n71. This work is licensed under CC BY 4.0. To view a copy of this license, visit https://creativecommons.org/licenses/by/4.0/

## Eligibility criteria

Definition and application of inclusion and exclusion criteria to maintain methodological rigor and ensure that each study addressed our research question were applied.

### Inclusion criterion

[1 Studies including patients diagnosed with Alzheimer’s disease (AD) according to established clinical or research diagnostic criteria.

1. Eligible studies investigated deep brain stimulation (DBS) targeting Fornix
2. Studies which in comparison group studied the conservative treatments or DBS in Other Areas
3. Studies which focused on Cognition as a primary outcome
4. studies that reported additional motor, neuropsychiatric, or quality-of-life outcomes if they provided extractable data on tremor randomized controlled trials (RCTs), quasi-randomized trials, and prospective or retrospective cohort studies were accepted. Limitation of time and language were not applied.

### Exclusion criterion

1. case reports were excluded. Also review articles, systematic reviews, meta-analyses, editorials, letters, and conference abstracts without complete data were excluded.
2. Animal studies.
3. studies with populations other than Alzheimer’s Disease, such as dystonia, essential tremor, or psychiatric conditions.
4. studies that only investigated DBS targets other than the Fornix
5. studies that lacked Cognition

Any disagreements until they reached consensus were solved by discussion, and when they could not agree, a third reviewer (FF) mediated to finalize the decision.

### Data extraction

Two reviewers (AM,AK) independently extracted data from each included study using a standardized Excel spreadsheet specifically developed for this review which designed by FF. We designed the extraction sheet to capture detailed information on study characteristics, patient demographics, surgical parameters, Cogniive outcomes, and methodological quality.

For each study, bibliographic and general information (title, first author, year, country, funding, conflicts of interest), study characteristics (design, single- vs. multi-center, study period, inclusion/exclusion criteria), and sample details, including total and subgroup sizes (Fornix, other conventional treatments), mean age, male sex distribution, disease duration, were extracted.

For each study, cognitive outcome measures (e.g., ADAS-Cog, MMSE, CDR-SB, or other validated cognitive scales), timepoints of assessment, baseline and post-intervention mean and standard deviation (SD) scores, and pre–post changes were extracted for both DBS-fornix and control groups. If multiple follow-up periods were reported, the longest available timepoint within the first year post-surgery and additional timepoints relevant for longitudinal analyses were included. Where reported, effect sizes and statistical results (e.g., t-test, ANCOVA, mixed-effects model) along with adjustment variables (age, disease duration, baseline cognitive severity) and data handling approaches (e.g., complete-case, imputation, LOCF, intention-to-treat vs. per-protocol) were extracted.

Information on surgical characteristics, including surgical approach, target laterality, DBS system brand, and stimulation parameters (e.g., amplitude, pulse width, frequency), were extracted separately for the fornix and control groups. Data on stimulation duration, adjustment and programming protocols, timing of first programming, number and duration of programming sessions, and the specific follow-up schedule were collected.

Adverse events (AEs) were extracted, including the type, number, rate, severity, timing relative to surgery, and relationship to the surgical or stimulation procedure. Handling of AEs, reporting duration, and outcomes of AE management were also recorded. Information regarding comorbidities, concomitant treatments, and baseline medications was extracted for both groups when reported.

### Risk of Bias Assessment

Two reviewers (RZ, NS) independently assessed the risk of bias using the Joanna Briggs Institute tools (JBI) appropriate for each study design (Supplementary 3). They resolved any disagreements through discussion or by consulting a third reviewer (FF) , ensuring a consistent and rigorous evaluation.

### Meta-Analysis

Two cognitive endpoints were analyzed separately: 1. ADAS Cog (Alzheimer’s Disease Assessment Scale–Cognitive Subscale), 2. MMSE (Mini Mental State Examination). For each study, means ± standard deviations of outcome scores at baseline and follow up were extracted from the Excel file, notably from: Standardized meandifferences (SMD; Hedges’ g) were computed using the metacont() function ofthe meta R package, which applies pooled standard deviations and the HartungKnapp adjustment under a random effects (REML) model.

### Main meta analysis

Separate pooled analyses were conducted for ADAS Cog and MMSE.Both outcomes showed no significant difference between DBS and control arms, ADAS Cog: pooled SMD ≈ 0, τ² = 0, I² = 0 %, MMSE (k = 3 ): pooled SMD = +0.054 [−0.524 to 0.633], p = 0.72, τ² = 0 .Forest plots displayed the individual and pooled effects with 95 % Cis around the null line and no visible heterogeneity.

Publication bias (Funnel & Trim-and-Fill) Publication bias wasinspected visually using funnel plots and by Duval and Tweedie’s trim-and-fill procedure.The ADAS-Cog plot (from adas cog funnel plot.pdf) showed four studies symmetrically distributed around SMD ≈ 0 ( −1.5 to 0 to +0.5).Trim-and-fill imputed one possible missing study (k = 1); the adjusted pooled estimate remained unchanged (+0.054 [−0.524, 0.63 3]), indicating no meaningful bias.

### Subgroup analysis

To evaluate whether study design affected outcomes, analyses were stratified by Design (RCT vs observational) using byvar = Design in side the meta function (to avoid update.meta errors).For MMSE, observational studies (Sankar 2020, Ponce 2014) and RCT (Leoutsakos 2018) yielded comparable effects (+0.38 vs +0.12; p ≈ 0.54).Subgrouping did not reduce τ², confirming that design was not a significant moderator.

### Sensitivity analysis

Leave one out analysis (using metainf()) evaluated result stability. Removing any single study (left Leoutsakos, Ponce, or Sankar) did not materially ch ange the pooled SMD or p value (all p ≫ 0.6), indicating robustness and no individual outlier influence. Meta regression Exploratory meta regression (using metafor::rma()) was performed on study l evel covariates (Design, Follow up months, Mean Age).Due to collinearity b etween Design and Follow up, only univariate models were run.The Mean Age effect (from meta regresion adas cog.pdf, with points Ponce 2014, Leo utsakos 2018, Sankar 2020 ranging 60– 70 years) showed a virtually flat regression line (β ≈ −0.04, p = 0.33, τ² = 0). No covariate significantly explained between study variance.

## Result

Ten studies included in this systematic review had multiple study designs. Three of them included data from a randomized, Phase IIb trial (the ADvance Trial, N = 42 mild AD) ^6–8^. There was a study with a retrospective cohort design, which compared DBS-fornix with conservative treatment ^4^. There were also three studies that were in the Phase I or feasibility stages^9–11^ a prospective case study focusing on five patients with severe AD ^12^, and several secondary or post-hoc analyses of larger trial datasets^8,13^. Most of the studies used data from the ADvance randomized controlled trial cohort ^6–8^, or merged data from ADvance with the earlier Toronto-based pilot trial to increase statistical power for specific analysis ^13^.

The sample sizes of these studies range from a single patient for a feasibility study ^9^to 58 patients in retrospective cohort design^13^. The target of the all of these studies was fornix while one retrospective study used DBS simultaneously in fornix and the nucleus basalis of Meynert (NBM) in severe AD^4^. The population of these studies mainly encompass the elder patients with mean ages ranging from 59.05±6.45 years in the DBS group for severe AD ^4^to 68.5±7.9 years in the mild AD cohorts^13^. The follow-up duration but was relatively heterogeneous between the studies ranging from the early post-operative evaluation (1.5-3 months)^7,12,13^ to long-term follow-up of one year^6,9,11^ and two years ^8^.(Table. 1)

**Table 1.**
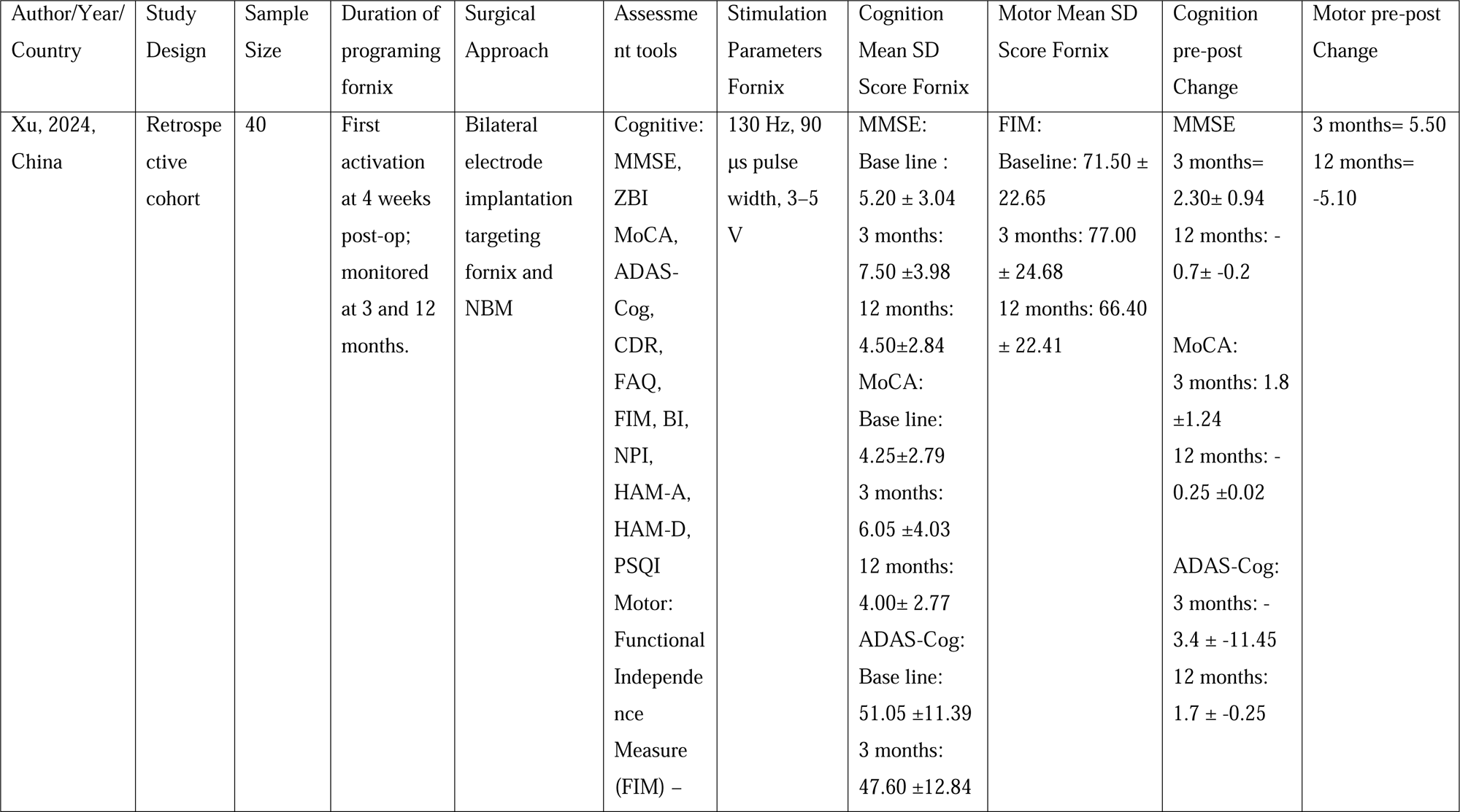
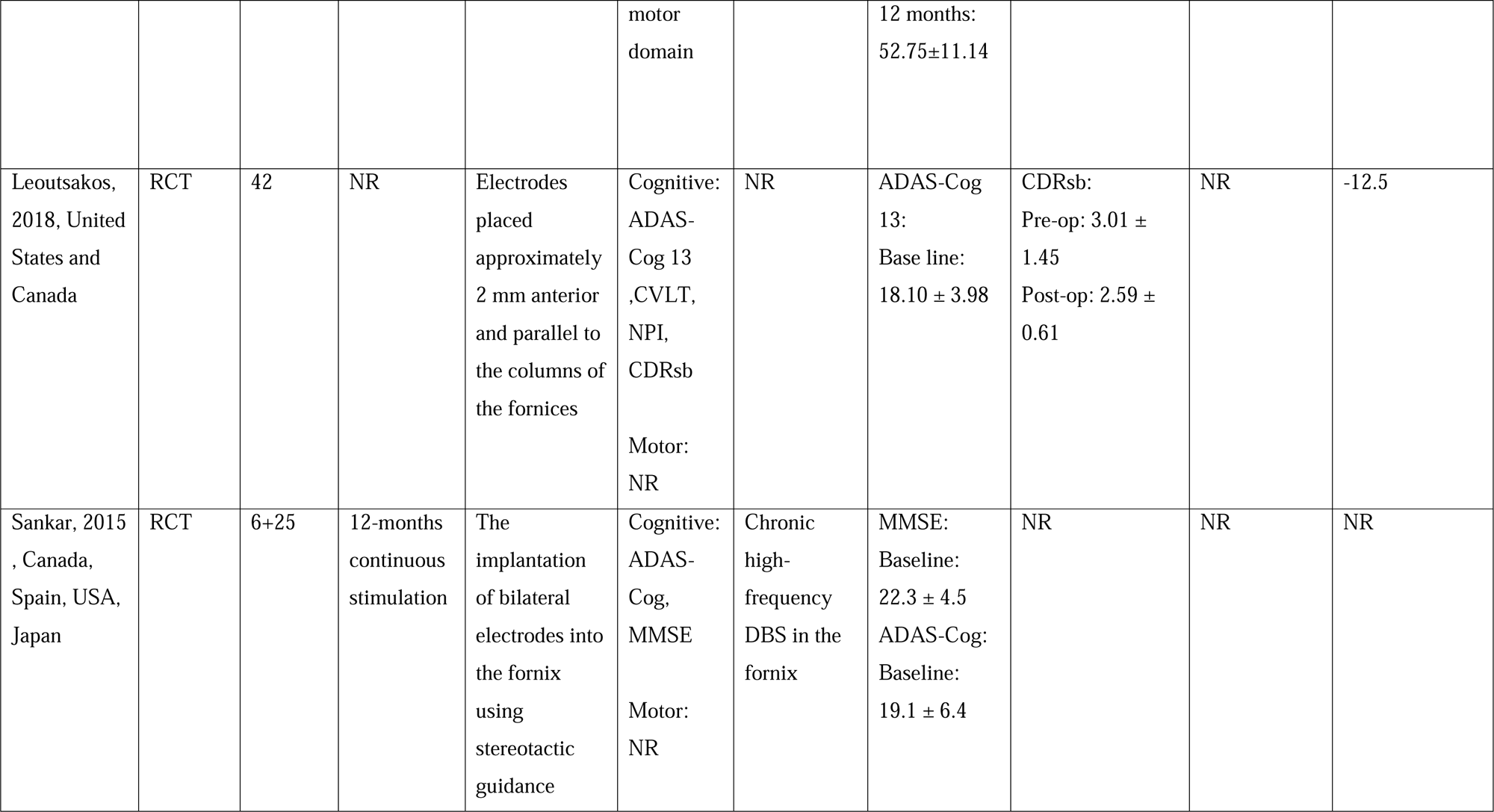
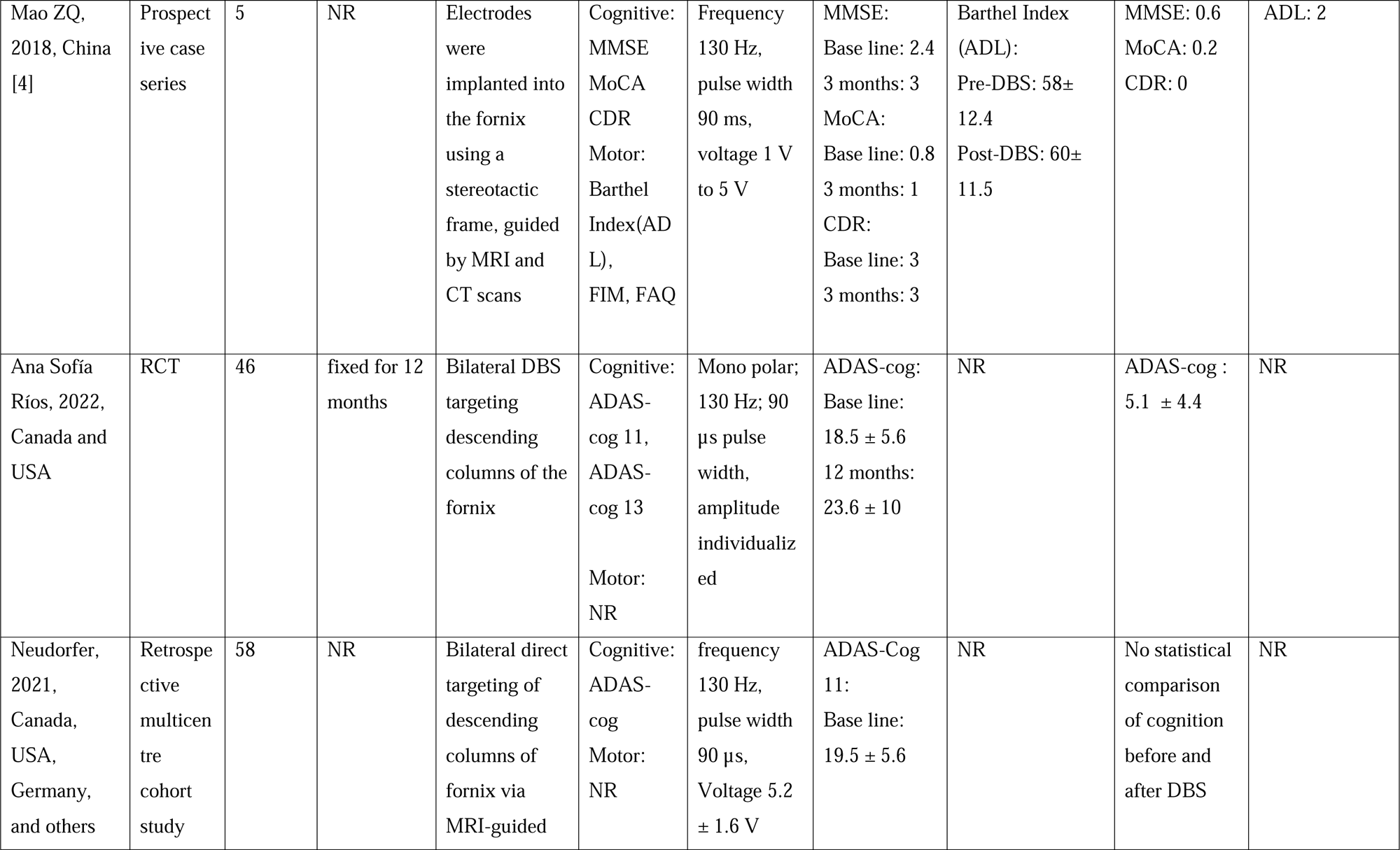
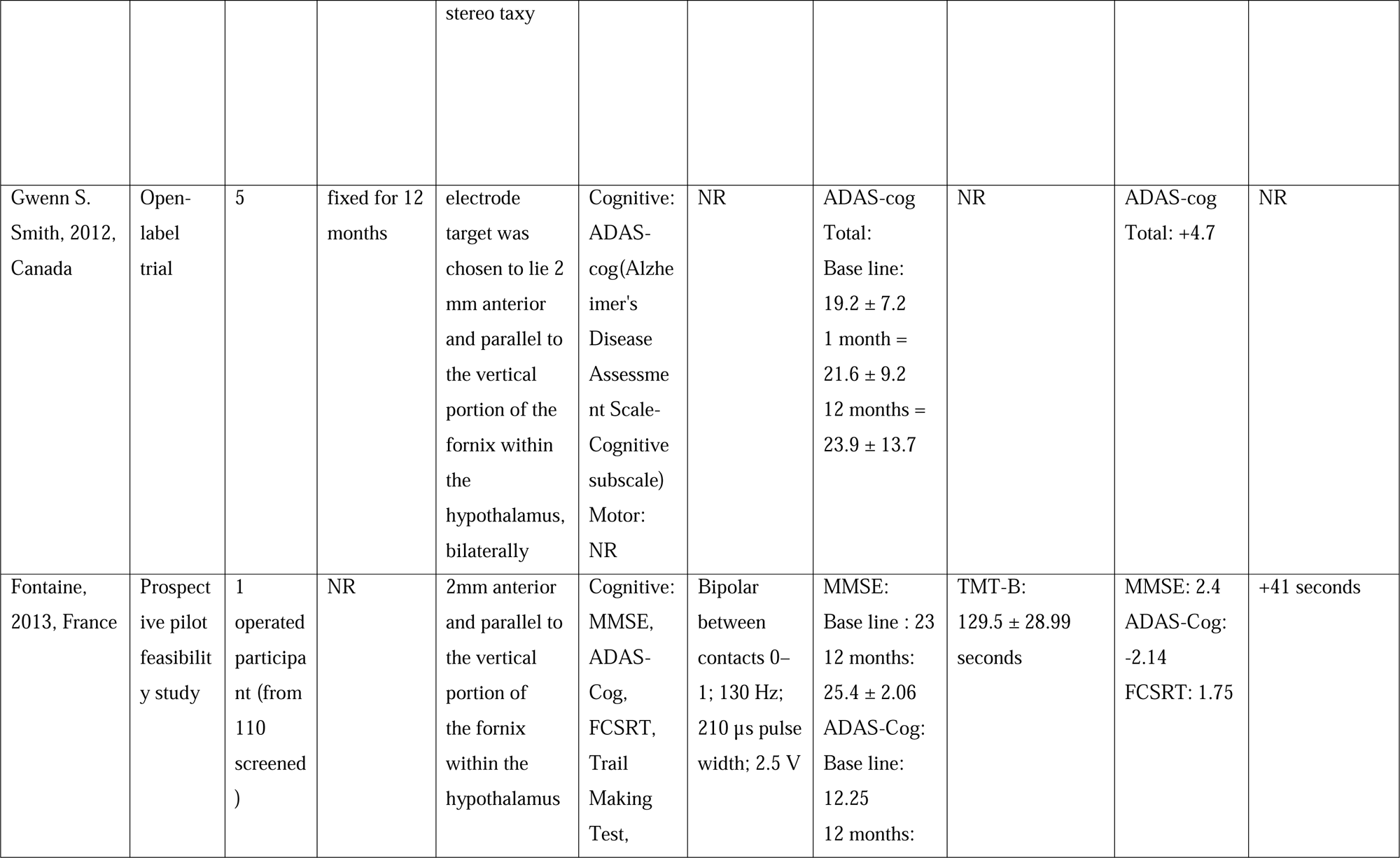
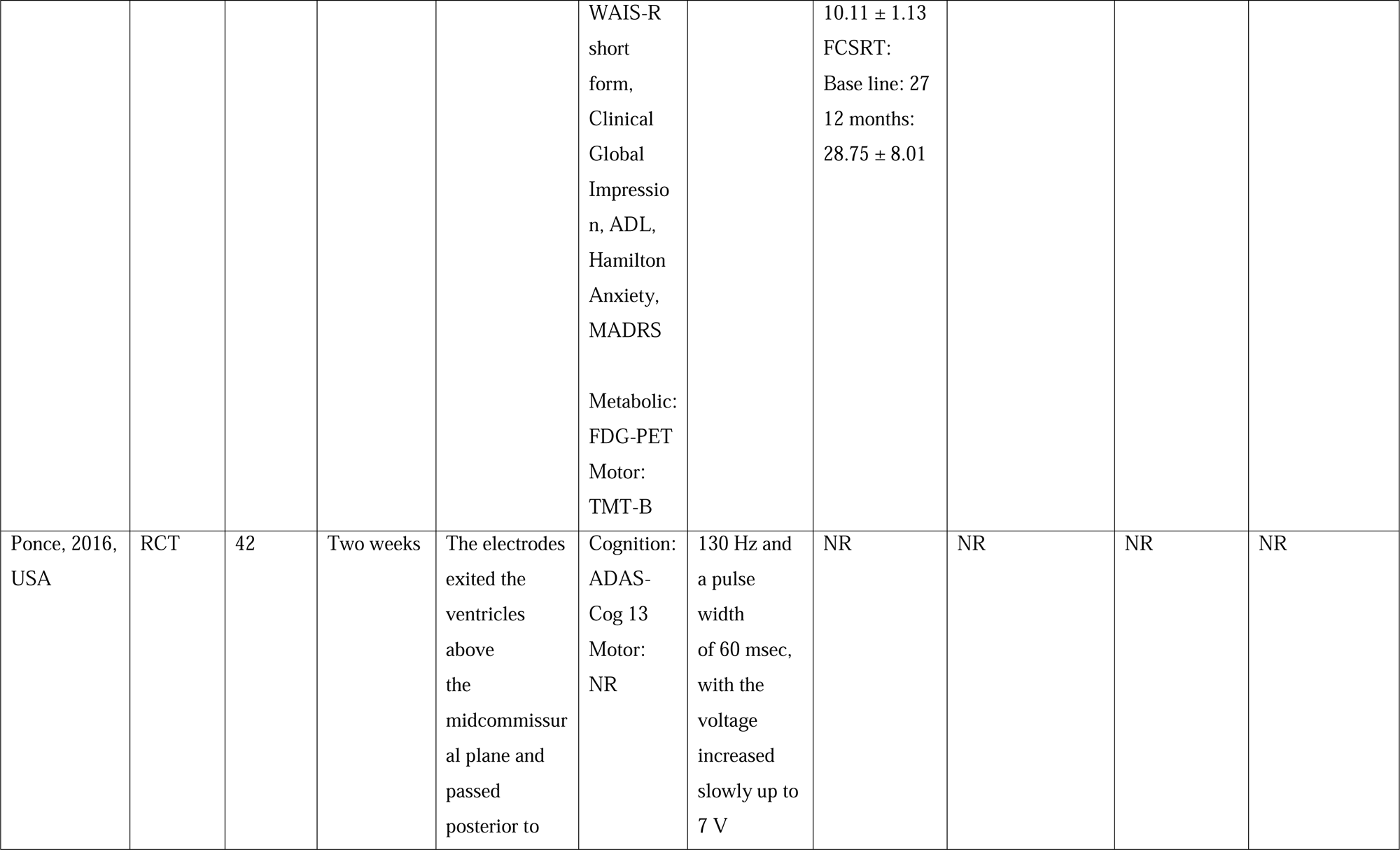
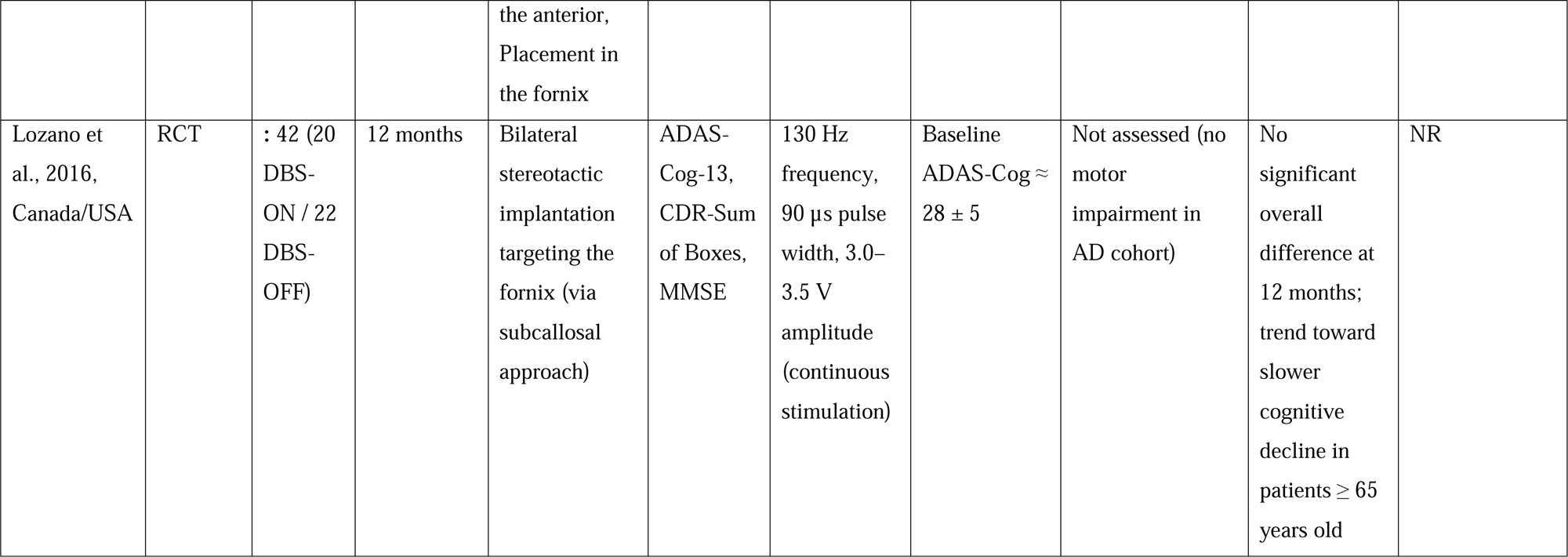
Summary of included studies evaluating deep brain stimulation (DBS) of the fornix in Alzheimer’s disease. The table presents study design, sample size, programming duration, surgical targeting approaches, cognitive and motor assessment tools, st

| Author/Year/<br>Country | Study<br>Design | Sample<br>Size | Duration of<br>programing<br>fornix | Surgical<br>Approach | Assessme<br>nt tools | Stimulation<br>Parameters<br>Fornix | Cognition<br>Mean SD<br>Score Fornix | Motor Mean SD<br>Score Fornix | Cognition<br>pre-post<br>Change | Motor pre-post<br>Change |
| --- | --- | --- | --- | --- | --- | --- | --- | --- | --- | --- |
| Xu, 2024,<br>China | Retrospe<br>ctive<br>cohort | 40 | First<br>activation<br>at 4 weeks<br>post-op;<br>monitored<br>at 3 and 12<br>months. | Bilateral<br>electrode<br>implantation<br>targeting<br>fornix and<br>NBM | Cognitive:<br>MMSE,<br>ZBI<br>MoCA,<br>ADAS-<br>Cog,<br>CDR,<br>FAQ,<br>FIM, BI,<br>NPI,<br>HAM-A,<br>HAM-D,<br>PSQI<br>Motor:<br>Functional<br>Independe<br>nce<br>Measure<br>(FIM) – | 130 Hz, 90<br>µs pulse<br>width, 3–5<br>V | MMSE:<br>Base line :<br>5.20 ± 3.04<br>3 months:<br>7.50 ±3.98<br>12 months:<br>4.50±2.84<br>MoCA:<br>Base line:<br>4.25±2.79<br>3 months:<br>6.05 ±4.03<br>12 months:<br>4.00± 2.77<br>ADAS-Cog:<br>Base line:<br>51.05 ±11.39<br>3 months:<br>47.60 ±12.84 | FIM:<br>Baseline: 71.50 ±<br>22.65<br>3 months: 77.00<br>± 24.68<br>12 months: 66.40<br>± 22.41 | MMSE<br>3 months=<br>2.30± 0.94<br>12 months: -<br>0.7± -0.2<br>MoCA:<br>3 months: 1.8<br>±1.24<br>12 months: -<br>0.25 ±0.02<br>ADAS-Cog:<br>3 months: -<br>3.4 ± -11.45<br>12 months:<br>1.7 ± -0.25 | 3 months= 5.50<br>12 months=<br>-5.10 |
|  |  |  |  |  | motor domain |  | 12 months:<br>52.75±11.14 |  |  |  |
| Leoutsakos, 2018, United States and Canada | RCT | 42 | NR | Electrodes placed approximately 2 mm anterior and parallel to the columns of the fornices | Cognitive:<br>ADAS-Cog 13<br>,CVLT,<br>NPI,<br>CDRsb<br>Motor:<br>NR | NR | ADAS-Cog 13:<br>Base line:<br>18.10 ± 3.98 | CDRsb:<br>Pre-op: 3.01 ± 1.45<br>Post-op: 2.59 ± 0.61 | NR | -12.5 |
| Sankar, 2015 , Canada, Spain, USA, Japan | RCT | 6+25 | 12-months continuous stimulation | The implantation of bilateral electrodes into the fornix using stereotactic guidance | Cognitive:<br>ADAS-Cog,<br>MMSE<br>Motor:<br>NR | Chronic high-frequency DBS in the fornix | MMSE:<br>Baseline:<br>22.3 ± 4.5<br>ADAS-Cog:<br>Baseline:<br>19.1 ± 6.4 | NR | NR | NR |
| Mao ZQ, 2018, China [4] | Prospective case series | 5 | NR | Electrodes were implanted into the fornix using a stereotactic frame, guided by MRI and CT scans | Cognitive: MMSE MoCA CDR Motor: Barthel Index(ADL), FIM, FAQ | Frequency 130 Hz, pulse width 90 ms, voltage 1 V to 5 V | MMSE: Base line: 2.4 3 months: 3 MoCA: Base line: 0.8 3 months: 1 CDR: Base line: 3 3 months: 3 | Barthel Index (ADL): Pre-DBS: 58±12.4 Post-DBS: 60±11.5 | MMSE: 0.6 MoCA: 0.2 CDR: 0 | ADL: 2 |
| Ana Sofía Ríos, 2022, Canada and USA | RCT | 46 | fixed for 12 months | Bilateral DBS targeting descending columns of the fornix | Cognitive: ADAS-cog 11, ADAS-cog 13 Motor: NR | Mono polar; 130 Hz; 90 µs pulse width, amplitude individualized | ADAS-cog: Base line: 18.5 ± 5.6 12 months: 23.6 ± 10 | NR | ADAS-cog : 5.1 ± 4.4 | NR |
| Neudorfer, 2021, Canada, USA, Germany, and others | Retrospective multicentre cohort study | 58 | NR | Bilateral direct targeting of descending columns of fornix via MRI-guided stereo taxy | Cognitive: ADAS-cog Motor: NR | frequency 130 Hz, pulse width 90 µs, Voltage 5.2 ± 1.6 V | ADAS-Cog 11: Base line: 19.5 ± 5.6 | NR | No statistical comparison of cognition before and after DBS | NR |
| Gwenn S. Smith, 2012, Canada | Open-label trial | 5 | fixed for 12 months | electrode target was chosen to lie 2 mm anterior and parallel to the vertical portion of the fornix within the hypothalamus, bilaterally | Cognitive: ADAS-cog(Alzheimer's Disease Assessment Scale-Cognitive subscale)<br>Motor: NR | NR | ADAS-cog<br>Total:<br>Base line:<br>$19.2 \pm 7.2$<br>1 month =<br>$21.6 \pm 9.2$<br>12 months =<br>$23.9 \pm 13.7$ | NR | ADAS-cog<br>Total: +4.7 | NR |
| Fontaine, 2013, France | Prospective pilot feasibility study | 1 operated participant (from 110 screened ) | NR | 2mm anterior and parallel to the vertical portion of the fornix within the hypothalamus | Cognitive: MMSE, ADAS-Cog, FCSRT, Trail Making Test, | Bipolar between contacts 0–1; 130 Hz; 210 μs pulse width; 2.5 V | MMSE:<br>Base line : 23<br>12 months: $25.4 \pm 2.06$<br>ADAS-Cog:<br>Base line: 12.25<br>12 months: | TMT-B: $129.5 \pm 28.99$ seconds | MMSE: 2.4<br>ADAS-Cog: -2.14<br>FCSRT: 1.75 | +41 seconds |
| | | | | | WAIS-R<br>short<br>form,<br>Clinical<br>Global<br>Impressio<br>n, ADL,<br>Hamilton<br>Anxiety,<br>MADRS<br>Metabolic:<br>FDG-PET<br>Motor:<br>TMT-B | | $10.11 \pm 1.13$<br>FCSRT:<br>Base line: 27<br>12 months:<br>$28.75 \pm 8.01$ | | | |
| Ponce, 2016,<br>USA | RCT | 42 | Two weeks | The electrodes<br>exited the<br>ventricles<br>above<br>the<br>midcommissur<br>al plane and<br>passed<br>posterior to the anterior,<br>Placement in<br>the fornix | Cognition:<br>ADAS-Cog 13<br>Motor:<br>NR | 130 Hz and<br>a pulse<br>width<br>of 60 msec,<br>with the<br>voltage<br>increased<br>slowly up to<br>7 V | NR | NR | NR | NR |
| Lozano et<br>al., 2016,<br>Canada/USA | RCT | : 42 (20<br>DBS-ON / 22<br>DBS-OFF) | 12 months | Bilateral<br>stereotactic<br>implantation<br>targeting the<br>fornix (via<br>subcallosal<br>approach) | ADAS-Cog-13,<br>CDR-Sum<br>of Boxes,<br>MMSE | 130 Hz<br>frequency,<br>90 μs pulse<br>width, 3.0–<br>3.5 V<br>amplitude<br>(continuous<br>stimulation) | Baseline<br>ADAS-Cog ≈<br>28 ± 5 | Not assessed (no<br>motor<br>impairment in<br>AD cohort) | No<br>significant<br>overall<br>difference at<br>12 months;<br>trend toward<br>slower<br>cognitive<br>decline in<br>patients ≥ 65<br>years old | NR |

### Cognition

The cognitive outcome of the DBS in fornix for the AD patients in these studies shows a heterogenus pattern which can be affected from multiple parameters including the severity and age of the patients. The cognitive function mainly measured by Alzheimer’s Disease Assessment Scale–Cognitive subscale (ADAS-Cog; scores reflecting worsening performance), the Mini-Mental State Examination (MMSE), and the Montreal Cognitive Assessment (MoCA) in these studies.

In Advance trial (N=42) DBS targeting of fornix did not significantly change he primary cognitive outcome (ADAS-Cog 13 and Clinical Dementia Rating Sum of Boxes [CDR-SB]) between the on-population and off-population during the first 12 months after the surgery^6^ in two year follow-up of the same patients also there was no significant difference between the treatment arms in cognitive outcomes (ADAS-Cog 13, CDRsb, California Verbal Learning Test [CVLT]) in either Phase 1 (0–12 months) or Phase 2 (12–24 months)^8^, but interestingly in posthoc sub-group analysis there was a difference in older patients (elder than 65 ) and younger patients ;In elder paricipants in 12 month follow-up there was a trend which shows that there was a significantly higher clinical outcomes in DBS-fornix in comparison with sham stimulation group^6^. The 2-year analysis reinforced this finding, noting that older participants in the early-on arm showed less worsening in Phase 1 compared to the delayed-on arm, and CDRsb scores deteriorated less in patients receiving DBS throughout the 24 months^8^. On the other hand for the younger patients it seems that DBS –fornix cause worsening (age<65,N=12) when the patients receiving he ON-DBS;ADA-Cog-13 score in this group of the patients decreased by 18.7±4.1 points which is greater than the OFF-DBS group (8.3±4.5), on the other cognitive scores like CDR-SB they find the same pattern which young patients detoriate larger in ON-DBS group (4.0 ± 0.7 ON vs 0.5 ± 0.5 OFF; p<0.001).

The results in the studies which focused on the sever-AD patients were different from the results described before, there was an study which focused on the 5 sever-AD patients receiving he DBS-Fornix for 1.5-3 monhes showed that all of the cognitive assesment scores improved except for CDR which remained unchanged in all cases ^12^.In this study the largest change in magnitude of scores occurred in MMSE and MoCA scores, beside these two the partial improvement also reported in mental state, language, long-term memory, and calculations^12^.

There was also another study which investigate the individuals with severe-AD retrospectively in cohort of 20 patients in comparison with 20 patients in control group (it is noteworthy to keep in mind that this study stimulated both fornix and nucleus basalis of Meynert), they showed that there was no significant difference between the DBS and control group in the baseline but the DBS group showed larger improvement in some cognitive scores in some time points, the MMSE scores at three months follow-up after surgery (P=0.002) were higher than the control and the MoCA scores at three months (P=0.003) and 12 months (P=0.010) after surgery had larger improvement in DBS group.

### Structural and Metabolic Correlates in Mild AD

Some of the studies used other metrics than the cognitive tasks to evaluate the efficacy of the fornix-DBS; one of the studies ^11^used the hippocampal volume to evaluate the efficacy, in their srudy 2 of the patients showed bilateral increase in the volume of the hippocampus after 12 month of fornix-DBS, in their studies these two patients also sgowed the best preservation of the cognitive function during the follow-up. Two other studies measured the glucose metabolism in the areas of brain that affected by AD to evaluate the fornix-DBS effect on these areas ^6,10^, in their studies in the one-year follow-up they found significant increase mainly in two networks frontal-temporal-parietal-striatal-thalamic network and a frontal-temporal-parietal-occipital-hippocampal network, Both a higher baseline metabolism (prior to DBS) and an increase in metabolism after 1 year of DBS in regions like the anterior cingulate, precuneus, and temporal gyri correlated with less decline or improvement in cognitive function ^10^. in Advance trial, the elder patients which (age>65) gets the fornix-DBS showed larger increase in the glucose metabolism in comparison with younger patients in 6 month follow-up.

### Meta-Analysis of the cognitive outcomes (ADAS-Cog)

From 10 included studies 4 of them reported data for cognitive outcome which assessed by Alzheimer’s Disease Assessment Scale–Cognitive Subscale (ADAS-Cog): Xu et al. (2015), Sankar et al. (2020), Ponce et al. (2014), and Leoutsakos et al. (2018). In total this 4 studies encompass 89 patients, 26 for observational study to 63 for RCT, the average age of the patients in these studies ranged from 60 to 70 years, a all of the 4 sudies reported follow-up duration of about 12 months. DBS electrodes implanted bilateraly in fornix region, the design of the observational studies was open-label design on the small population of the patients (Xu 2015; Sankar 2020), but the RCTs compared the DBS ON condition with conservative standard treatment. (Ponce 2014 and Leoutsakos 2018)

### Quantitative Synthesis

Pooled analysis of the studies showed no statistically significant effect of the DBS-f on ADAS-Cog scores in comparison with control group, the heterogeneity across studies was relatively low (τ² =0.14, I² = 57.2 %), with indicates that the result of the studies were consistent.(Fig 2)

**Figure 2.**
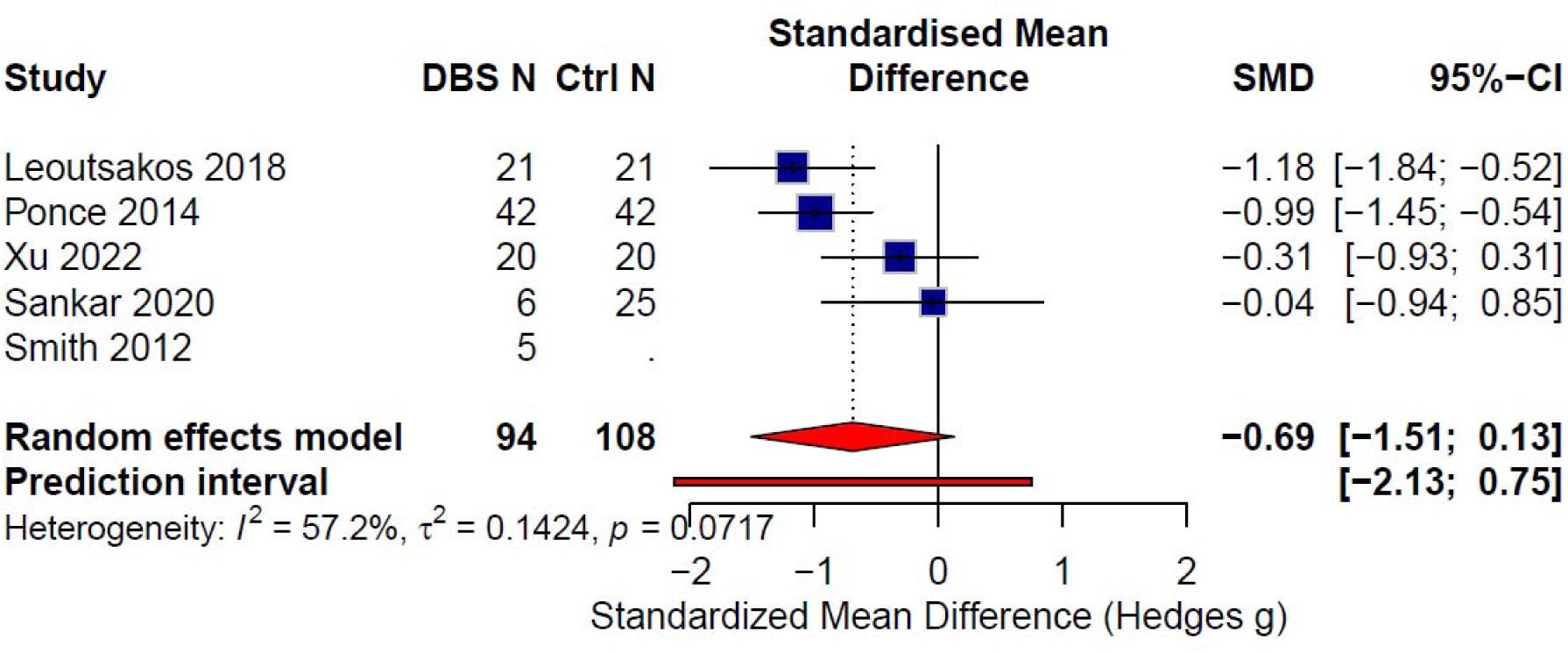
Pooled effect of DBS-f on ADAS-Cog scores.Meta-analysis showed no significant difference between DBS-f and control groups, with low heterogeneity (τ² = 0.14, I² = 57.2%).

### Subgroup Analyses

A subgroup comparison based on study design (observational vs. RCT) revealed that there was no significant difference in treatment effect (χ²(1) = 1.74, p = 0.19) across study designs. Thus, cognitive outcomes following fornix DBS were similar in both observational and randomized designs. (Fig.3)

**Figure 3.**
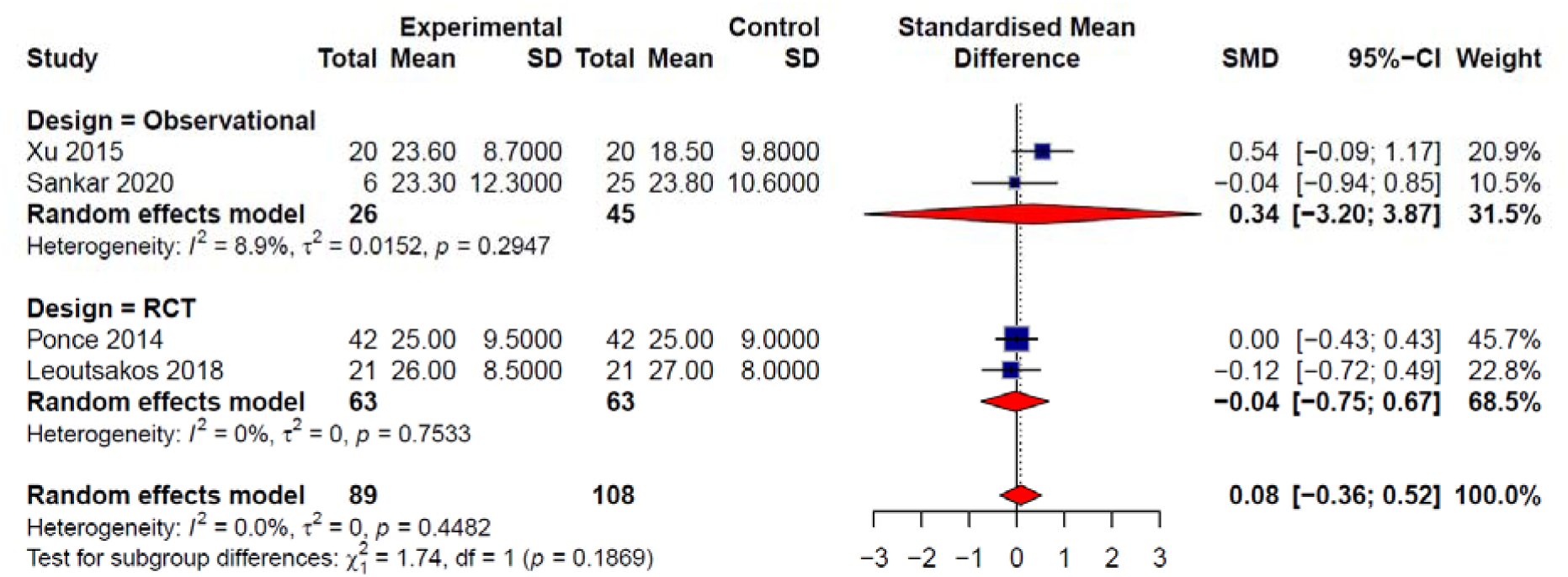
Subgroup analysis by study design.Comparison of observational studies and RCTs showed no significant difference in treatment effect (χ²(1) = 1.74, p = 0.19), indicating similar cognitive outcomes across designs.

### Publication Bias

Visual inspection of the funnel plot showed the four studies symmetrically distributed around the null line (SMD ≈ 0, range −1.5 to +0.5). The Duval and Tweedie trim-and-fill method yielded k = 0, suggesting no missing studies and therefore no evidence of publication bias. The adjusted pooled estimate (SMD = 0.08 [−0.36, 0.52]) was identical to the unadjusted value.(Fig 4)

**Figure 4.**
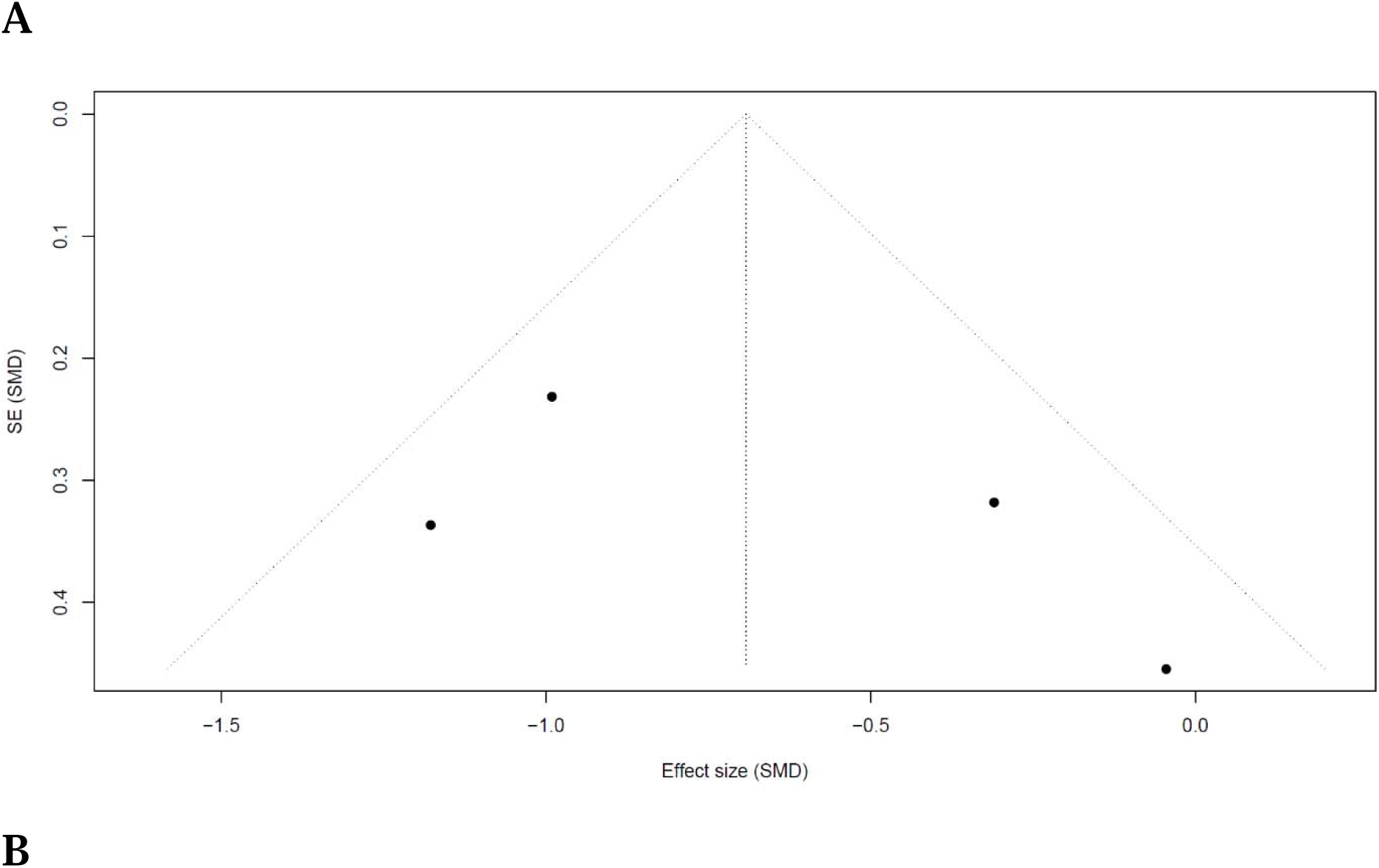
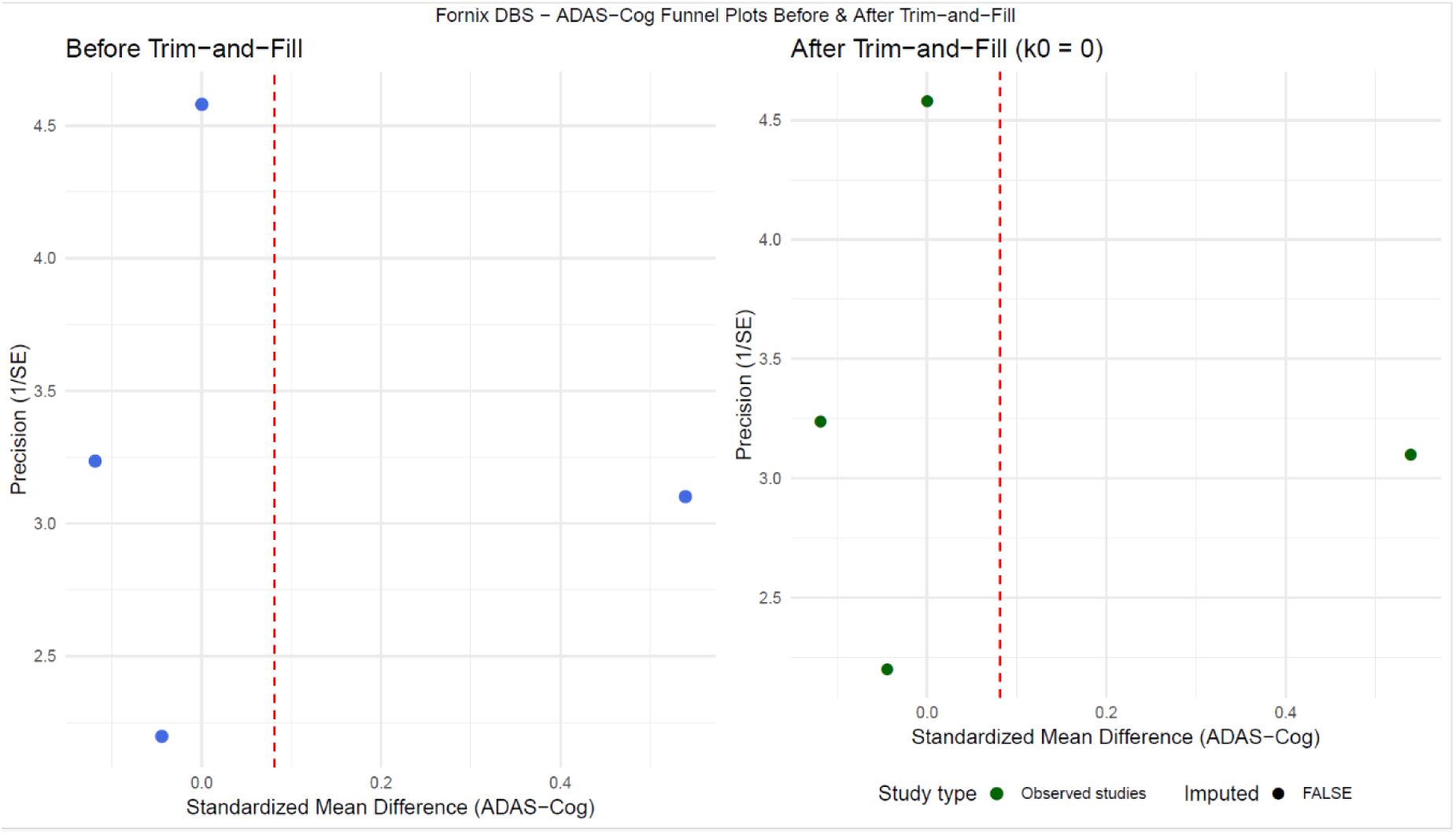
Assessment of publication bias.(a) Funnel plot showing symmetric distribution of studies around the null effect, indicating no visual evidence of publication bias. (b) Duval and Tweedie trim-and-fill analysis identified no missing studies (k[ = 0), and the adjusted pooled effect (SMD = 0.08 [−0.36, 0.52]) was unchanged.

### Meta-Regression

A univariate meta-regression of study-level mean age (60–70 years) against effect size demonstrated no association (β ≈ −0.04, p = 0.33, τ² = 0), indicating that age did not explain between-study variation. Other potential moderators (follow-up duration, study design) were collinear and not retained. (Fig.5)

**Figure 5.**
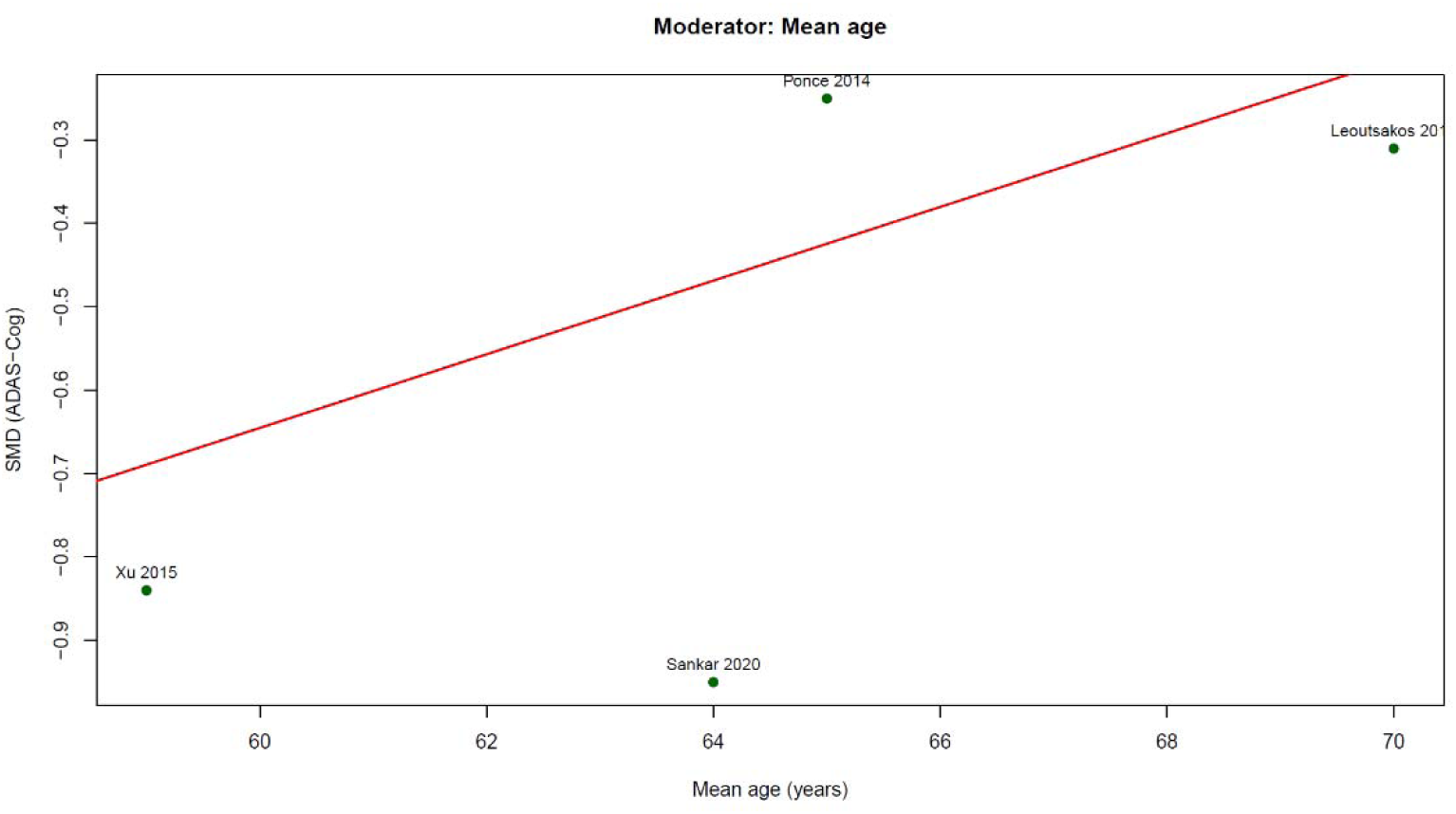
Meta-regression analysis.Univariate meta-regression showed no association between mean age and effect size (β ≈ −0.04, p = 0.33), indicating that age did not account for between-study variation.

### Meta-Analysis of the cognitive outcomes (MMSE)

We included three studies in quantitative meta-analysis of the Mini-Mental State Examination (MMSE) outcome; Sankar et al. (2020), Ponce et al. (2014), and Leoutsakos et al. (2018). Two of the studies - Ponce (2014) and Leoutsakos (2018)- were RCTs, on the ADvance study, which Leoutsakos (2018) represents results from follow-up phase of RCT, and the third one was an observational, open labeled study Sankar (2020). Sample sizes of the included studies ranged from 12 to 33 patients in each arm, with 21 in control group and 33 in DBS-f patients, the average age in studies were between 63 and 65 years and the duration was approximately about 12 months in all patients.

### Overall Pooled Effect

The pooled analysis showed that there was no significant difference in change in MMSE between DBS-f and control group, the standardized mean difference (SMD) was +0.05 [−0.52, +0.63], p = 0.72. we performed the heterogeneity analysis which demonstrated that between study heterogeneity is negligible (τ² = 0; I² = 0 %; χ² = 0.59, p = 0.74), which showed that the results in studies were aligned and the null effect were consistent in all studies. (Fig. 6)

**Figure 6.**
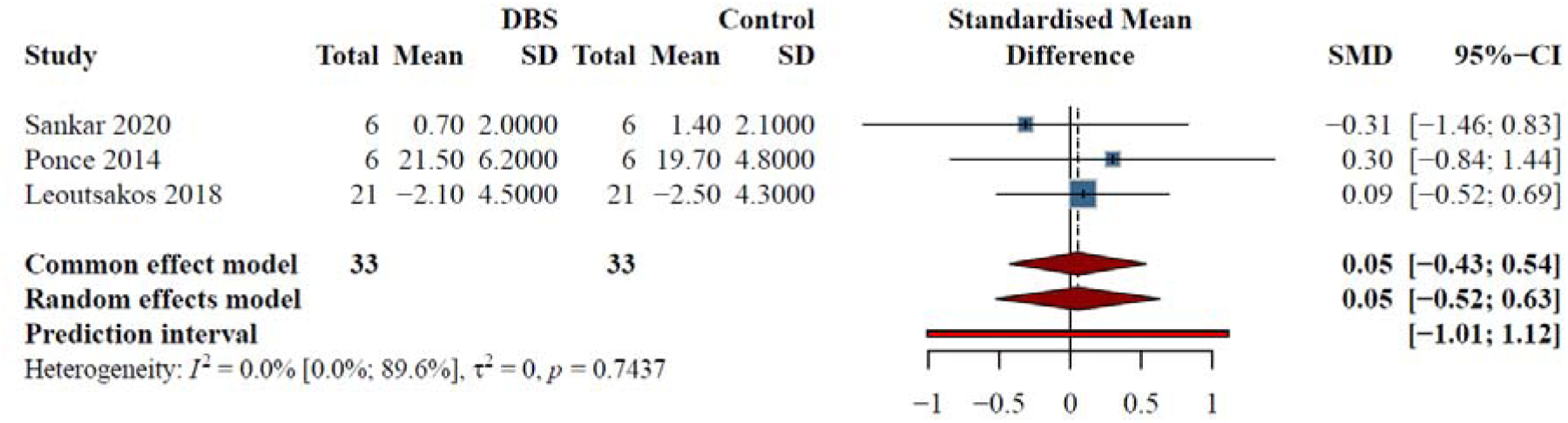
Pooled analysis of change in MMSE scores between DBS-f and control groups. The standardized mean difference (SMD) showed no significant effect (+0.05 [−0.52, +0.63], p = 0.72). Heterogeneity across studies was negligible (τ² = 0; I² = 0%; χ² = 0.59, p = 0.74), indicating consistent results.

### Subgroup Analyses

A subgroup comparison based on study design (observational vs randomized) performed and revealed no significant difference under either model: Common-effects: χ²(1) = 0.04, p = 0.85, Random-effects: χ²(1) = 0.05, p = 0.82 Thus, whether DBS was evaluated in a controlled or observational framework did not influence MMSE outcomes.(Fig. 7)

**Figure 7.**
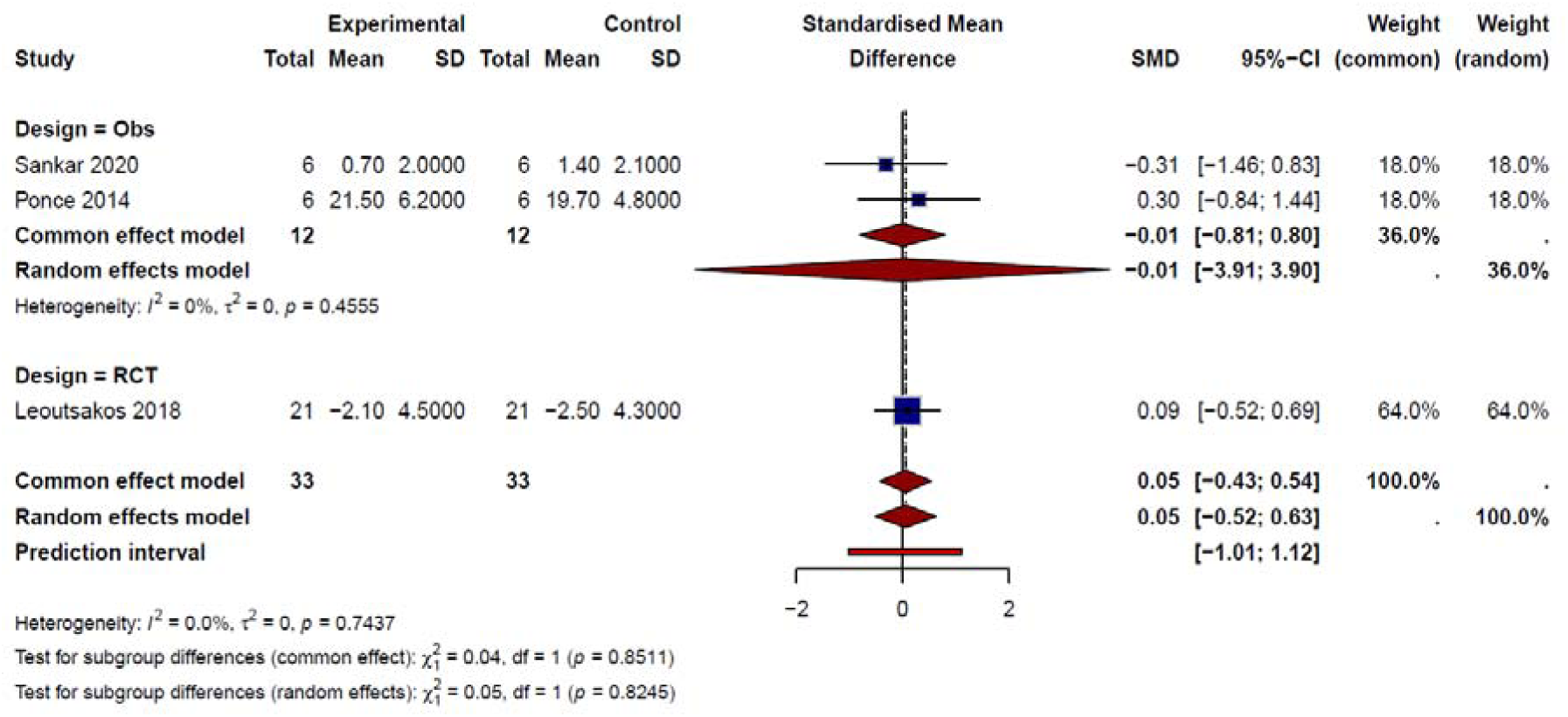
Subgroup analysis of MMSE change by study design (observational vs. randomized) showing no significant difference under common-effects (χ² = 0.04, p = 0.85) or random-effects (χ² = 0.05, p = 0.82) models, indicating study design did not affect outcomes.

### Publication Bias

Visual inspection of the funnel plot showed a symmetric distribution of studies around the null line (SMD ≈ 0). Application of the Duval and Tweedie trim-and-fill procedure yielded k = 0, implying no imputed missing studies and therefore no evidence of publication bias. The adjusted pooled estimate (SMD ≈ +0.05) was identical to the unadjusted model, underscoring the stability of the null result.

Qualitatively, all points were distributed near the origin with balanced dispersion, satisfying Egger’s visual criterion for funnel-plot symmetry. (Fig. 8)

**Figure 8.**
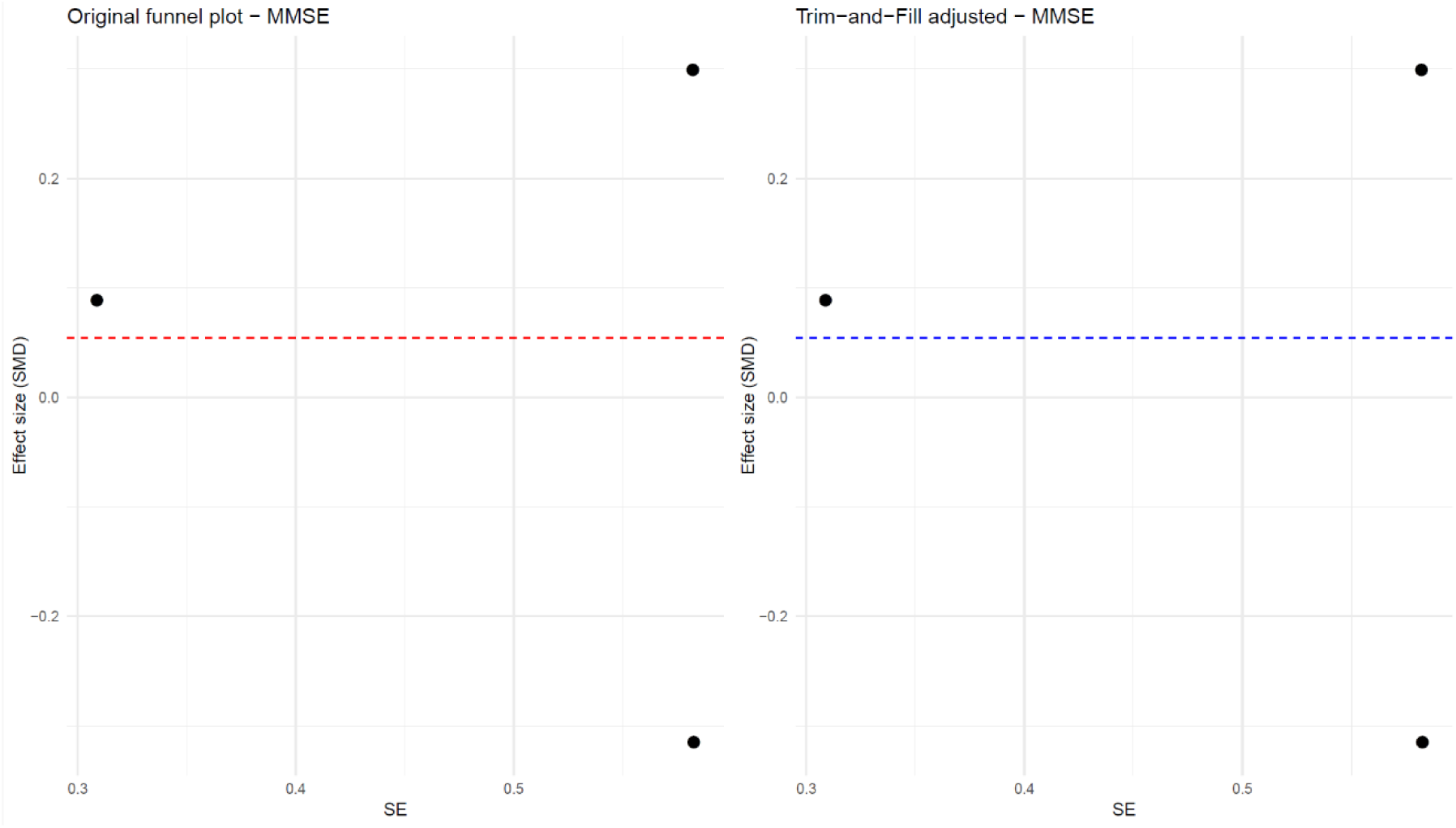
Funnel plot assessing publication bias in MMSE outcomes. Studies were symmetrically distributed around the null (SMD ≈ 0). Trim-and-fill analysis imputed no missing studies (k[ = 0), and the adjusted pooled estimate remained +0.05, indicating no evidence of publication bias.

### Meta-Regression

A univariate meta-regression using mean age (63–65 years) as the covariate indicated no significant association with effect size (β ≈ −0.04, p = 0.33; τ² = 0). Other potential moderators (e.g., follow-up duration, study design) were collinear and therefore excluded.(Fig. 9)

**Figure 9.**
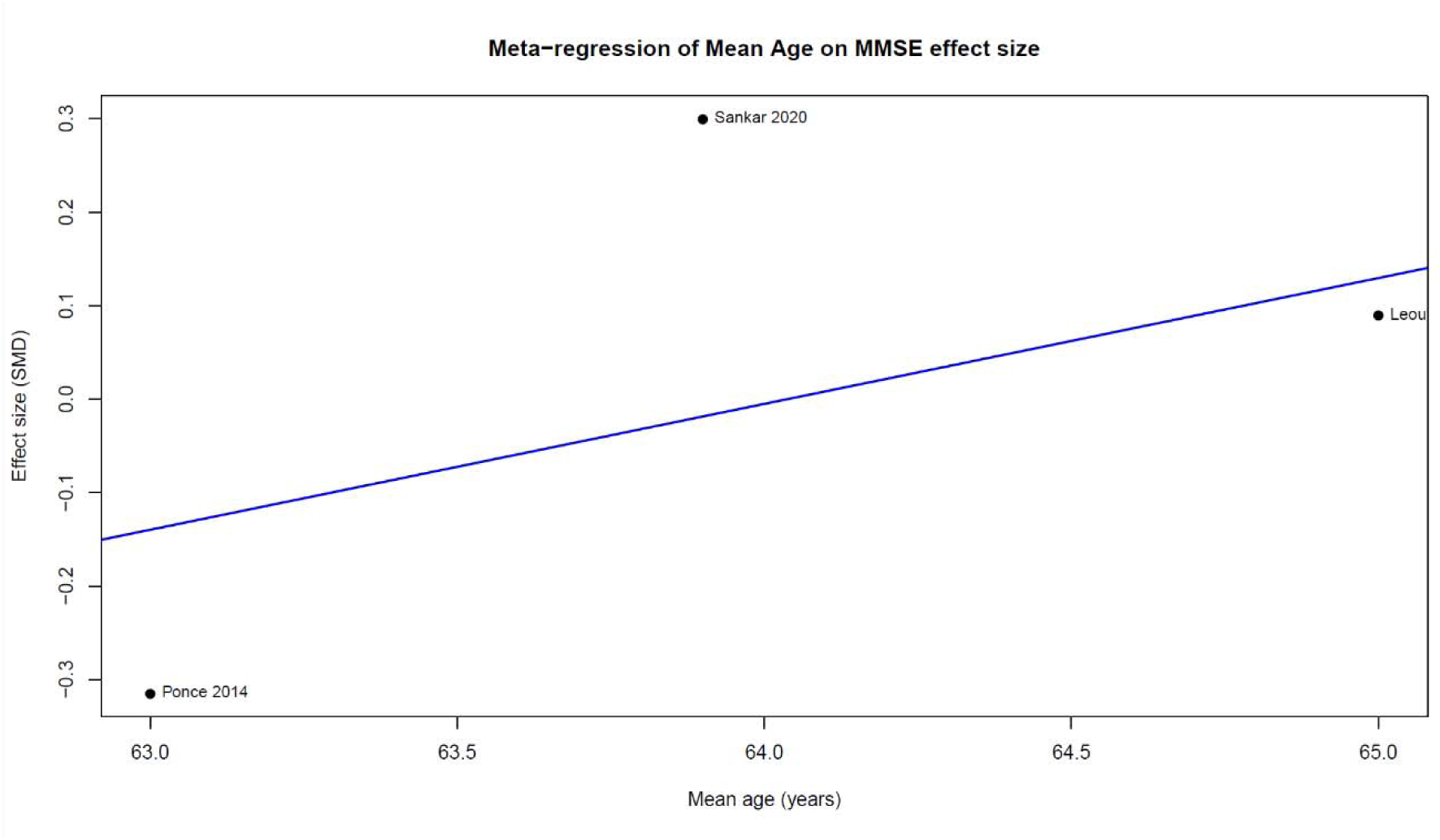
Univariate meta-regression of MMSE change by mean age (63–65 years) showing no significant association with effect size (β ≈ −0.04, p = 0.33; τ² = 0). Other potential moderators were excluded due to collinearity.

### Motor

The measurement tools for the motor outcomes in the studies include activities of daily living (ADL) and functional independence, such as the Functional Independence Measure (FIM) and the Barthel Index (BI). The FIM tool has two parts for cognition(35 points) and motor(91 points), which lower FMI score shows the lower ability in self-care.

In a retrospective cohort study which compare the DBS with conservative treatments, showed a notable improvement in FIM score for the DBS group. the baseline FIM score was relatively similar in two groups; DBS group averaging 71.50±22.65 points and the control group averaging 69.55±26.18 points, Raw FIM scores measured three months (77.00±24.68 in DBS vs. 72.20±25.98 in control) and 12 months (66.40±22.41 in DBS vs. 67.20±25.3 in control) post-treatment did not show a statistically significant difference between the two groups (three months: p=0.323; 12 months: p=0.968) ^4^.But when we focus on the magnitude of the improvement; the DBS group demonstrated significantly greater improvements in FIM scores three months after surgery compared to the control group (p < 0.05). This significant difference in improvement was not sustained at the 12-month follow-up.

In a smaller study which involved five patient with sever DBS-f no significant improvement observed in ADL in early post-surgery duration but the single patient analysis was mixed : one patient experienced a degeneration in basic ADL, indicated by the Barthel Index falling from 55 to 45, while another severe case showed increases in the Barthel Index and FIM scores to different degrees.

### Surgical approach employed by the studies

Across the included studies, the surgical strategies employed was bilateral DBS targeting fornices in all included studies. None of the included studies reported unilateral DBS targeting fornices surgical approach. According to fornix’s anatomy; in 6 included studies implanted the electrodes of DBS in column of fornices ^7–10,13^.In 3 included studies didn’t determined the exact place of electrodes implantation in the fornices^4,11,14^.

### Deep Brain Stimulation Parameters

Among the included studies, 8 of them reported the exact amount of frequency, pulses and voltages that they used for DBS in their studies^4,7,9,11–14^. Four included studies used the same frequency and pulses (130 HZ, 90 µs pulses), only their was differences between voltage that they used, which Xu et al used 3-5 voltage, Mao ZQ et al used 1-5 voltage, Ríos et al didn’t report the exact amount of voltage, and Neudorfer et al used 5.2 ± 1.6 voltage^12–14^. Fontaine et al used 130 Hz frequency, 210 µs pulse width, and 2.5 voltage ^9^. Ponce et al used less pulses (60 msec), and more voltage (7 V) than Fontaine et al study, but the same frequency (130HZ) ^7,9^.

### Duration of programming fornix

Only 5 included studies described the time of programing for fornix throughout all included studies ^4,7,10,11,14^. And five remaining studies didn’t show the time of programing for fornix ^6,8,9,12,13^.Xu et al described their study by the First activation at 4 weeks and post-op monitorig at 3 and 12 months ^4^. Sankar et al recorded 12-months continuous stimulation in their project ^11^. Ríos et al and Smith et al in their research fixed the programing of fornix for 12 months ^7,10^. Ponce et al programmed the duration of their study less than other included studies for only Two weeks ^7^.

### Adverse Events

The adverse events after the DBS in fornix for the Alzheimers disease in included studies can be classified in Surgical and Device-Related Complications, Acute Stimulation-Induced Effects (Intraoperative/Programming), and General/Chronic Medical and Psychiatric Events. Generally the DBS surgery in fornix fo the patients with dementia considered safe and well-tolerated in most cases.

The surgical- and device-related adverse events usually occur around the time of surgery or within the first three months after the procedure. In the ADvance Trial (N = 42 patients with mild AD), 61.9% of patients experienced 64 adjudicated adverse events (AEs) related to the procedure, with seven serious adverse events (SAEs) reported in 11.9% of patients within 90 days ^7^.

The most frequent adverse effect was related to the surgical site (31.3%), followed by headache (23.4%) ^7^. Specific adverse events requiring return to the operating room included infections (in two patients) related to device implantation and one case of subdural hematoma^6,7^.

In a study involving patients with severe AD, 30% of participants (11 adverse events) reported AEs ^4^. Two patients experienced severe AEs due to chronic subdural hematoma, which were successfully treated with puncture and drainage. Mild surgical or general AEs in this severe AD cohort included inflammation at the pulse transmitter site, postoperative pain, swelling, and redness — all of which resolved with symptomatic treatment ^4^.

### Risk of Bias Assessment

The ten studies reviewed showed a low overall risk of bias. Nine studies had similar groups, controlled for confounders, and used valid methods to measure exposure. All studies used valid outcome assessments and ensured that participants did not have the outcome at baseline. Eight studies had a long enough follow-up period. About half of the studies clearly explained the extent of their follow-up and why some participants dropped out. Five studies clearly described their statistical methods, but the others did not provide enough detail. Only one study was judged high risk, and this was due to the way exposure was measured. Overall, the studies were methodologically strong, with most issues coming from unclear reporting rather than a high risk of bias.(Fig. 10)

**Figure 10.**
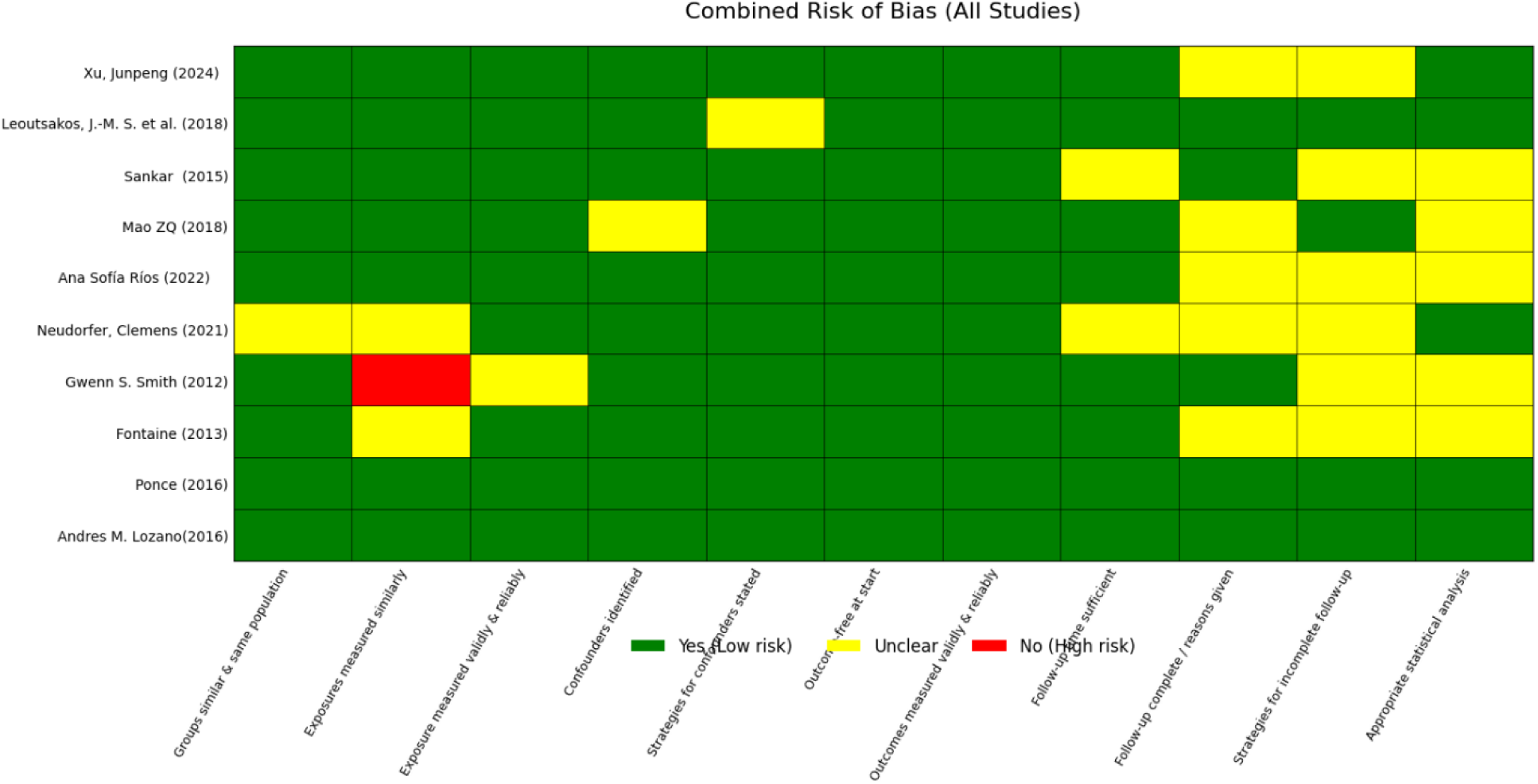
Risk-of-bias assessment of the ten included studies. Overall, studies demonstrated low risk of bias, with most concerns arising from unclear reporting rather than methodological flaws. One study was rated high risk due to exposure measurement.

## Discussion

### Main Findings

This systematic review and meta-analysis, synthesizing data from ten studies, including randomized controlled trials ^2,7,9^ and observational or retrospective cohorts^1,11^, shows that deep brain stimulation of the fornix (DBS-f) does not provide sustained cognitive improvement in Alzheimer’s disease during approximately 12 months of follow-up. in both major cognitive outcomes, ADAS-Cog and MMSE, the pooled effect sizes remained near to zero (ADAS-Cog: SMD = 0.08 [−0.36, 0.52], τ²=0.14, I²=57%; MMSE: SMD = 0.05 [−0.52, 0.63], τ²=0, I²=0), confirming a null effect despite heterogeneity in study design, disease severity, and stimulation parameters.

### Short-term vs. Long-term changes

Short-term improvements, particularly within the first three months, were reported primarily in severe AD cohorts^1,4,8^. Xu et al. observed MMSE increasing from 5.20 ± 3.04 to 7.50 ± 3.98 (+2.30, p<0.01) and MoCA improving by +1.8. However, these improvements consistently decrease under baseline after 12 months. Mao et al. reported improvement across MMSE subdomains except CDR. In contrast, multicenter RCTs ^2,9^ demonstrated no meaningful benefit at 12 months, with some showing even faster deterioration. This supports a biphasic pattern: transient early gains followed by progressive decline.

### Age and Disease Stage Effects

Age plays a role as a major modifier of treatment response. The ADvance trial ^2^found that patients older than 65 years old exhibited slower cognitive decline in the procedure, whereas younger patients showed accelerated deterioration. With this explanation, advanced degeneration in older patients may allow DBS to reactivate residual circuits, whereas stimulation in younger patients may disrupt relatively preserved networks. Severe-AD cohorts showed early improvements, which support the idea that disrupted network may increase DBS responsiveness.

### Mechanistic Correlates

Some physiological evidences confirm that DBS-f affects memory circuits, even when cognitive outcomes remain unchanged. Sankar et al. (2020) reported bilateral hippocampal volume increases after 12 months in two patients with best cognitive preservation^11^. PET studies (Smith et al., 2012 ; Lozano et al., 2016) demonstrated increased glucose metabolism in two major networks: a frontal-temporal-parietal-striatal-thalamic network and a frontal-temporal-parietal-occipital-hippocampal network. Greater baseline metabolism and increases in regions such as the anterior cingulate and precuneus correlated with less cognitive decline^6,7^. In the ADvance trial, older patients exhibited larger metabolic increases at six months.

### Meta-analysis Findings

Four studies provided ADAS-Cog data (Xu 2015; Sankar 2020; Ponce 2014; Leoutsakos 2018). Together they included 89 patients. The pooled effect showed no benefit (SMD = 0.08; τ²=0.14; I²=57.2%), with consistent results between observational and RCT designs. MMSE pooled effect was similarly null (SMD = 0.05; τ²=0; I²=0). Meta-regression demonstrated no association between age and effect size (β = −0.04, p=0.33). Trim-and-fill detected no missing studies (k =0), and funnel plots showed symmetry.

### Motor and Functional Outcomes

Motor outcomes were rarely assessed and generally reported in the setting of general functional scales. In a 20-patient severe-AD cohort^1^, reported that the FIM scores did not differ significantly at 3 or 12 months (p=0.323; p=0.968). However, FIM improvement support that DBS can show benefit at three months (p<0.05), but this benefit disappeared at 12 months. A smaller study (Mao et al., 2018 [4]) showed mixed single-patient outcomes: one case declined (Barthel 55→45), another improved. Overall, motor/ADL benefits appear transient and likely reflect attentional/arousal effects rather than motor-circuit modulation.

### Surgical Approach and Stimulation Parameters

All included studies used bilateral implantation. Six studies placed electrodes in the columns of the fornix^7–10,13^, while three did not specify precise placement [4,11,14]. No study used unilateral DBS. Parameter variability was considerable. Most studies used 130 Hz and near 90 µs pulse width, but voltages ranged from 1–7 V. Xu used 3–5 V; Mao 1–5 V; Fontaine 2.5 V with 210 µs; Ponce used 7 V with 60 µs. This heterogeneity results in inconsistent outcomes.

### Follow-Up Duration and Adaptation

Follow-up periods differ from two weeks to two years. Short intervals (e.g., Ponce, two weeks) showed no effect, whereas longer follow-up demonstrated the typical biphasic response: early improvement followed by^1,10,11^. This suggests transient network adaptation rather than lasting neuromodulation.

### Clinical Implications and Future Directions

In this review several clinical implications achieved. Although fornix-targeted DBS consistently engages memory-related circuits, this physiological activation does not translate into sustained cognitive improvement. Quantitative synthesis showed that mean differences in ADAS-Cog and MMSE were small and centered around zero, with low-to-moderate heterogeneity, indicating a consistent null effect across study designs and populations. This suggests that while the fornix remains metabolically responsive, DBS cannot overcome the progressive synaptic degeneration characteristic of Alzheimer’s disease.

One major implication concerns age-dependent variability in the ADvance trial. Older participants (≥65) exhibited modest slowing of decline under DBS, whereas younger individuals (<65) deteriorated more rapidly (Lozano, 2016; Leoutsakos, 2018). In the younger group, ADAS-Cog declined by 18.7 ± 4.1 points under DBS compared with 8.3 ± 4.5 in the sham condition. Similarly, CDR-SB worsened significantly faster (4.0 ± 0.7 ON vs 0.5 ± 0.5 OFF). These results can confirm that DBS-f does not have a uniform therapeutic effect across age groups. Older patients with extensive circuit degeneration may benefit from re-activation of remaining limbic fibers, while younger patients may experience disruption of endogenous hippocampal–fornical circuits.

Disease severity also shows related dichotomy. In severe AD, short-term improvements were frequently observed. For example, Mao (2018) reported that all measured cognitive domains except CDR improved in five severe-AD patients during 1.5–3 months of DBS. Another retrospective severe-AD cohort (Xu, 2024), using dual stimulation of the fornix and nucleus basalis of Meynert, reported significantly higher MMSE at three months (p=0.002) and higher MoCA at three months (p=0.003) and 12 months (p=0.010). These results suggest that in advanced disease stages, transient activation of residual limbic pathways can produce improvements. In contrast, mild-AD patients in controlled trials (Ponce, 2014; Leoutsakos, 2018) showed no meaningful benefit even at early timepoints, suggesting a limited therapeutic window for DBS-f.

Technical factors also have clinical implications. Studies differed substantially in stimulation parameters, ranging from 1–7 V, pulse widths from 60–210 µs, and frequencies from 40 to 130 Hz, and programming duration varied widely, from as short as two weeks (Ponce, 2014) to continuous stimulation over 12 months (Sankar, 2015; Ríos, 2016; Smith, 2012). This heterogeneity indicates that current parameter sets are not optimized for cognitive modulation, and almost extracted from DBS parameters in the setting of parkinsonism patients.

The meta-analysis strengthens these observations: neither subgroup analysis nor meta-regression showed any modifying effect of age or study design on cognitive outcomes, reinforcing the conclusion that the absence of measurable cognitive benefit is robust across methodological differences. Early improvements documented in open-label settings uniformly dissipated with longer follow-up, supporting the hypothesis that DBS-f primarily induces short-lived network activation rather than durable neurorestorative change.

Motor outcomes add further nuance. Despite occasional early improvements in functional indices, such as FIM improvement at three months in the DBS group (p<0.05) in the severe AD cohort (Xu, 2024), no sustained benefit was present at 12 months. A five-patient severe AD study reported mixed ADL outcomes, including decline in one patient’s Barthel Index (55→45). These patterns suggest that functional gains likely reflect transient increases in attention or arousal rather than genuine motor improvement.

Taken together, the totality of evidence suggests that DBS-f should not currently be considered a disease-modifying therapy for Alzheimer’s disease.

## Limitation

First, the included studies exhibited methodological heterogeneity: designs ranged from randomized controlled trials to retrospective cohorts and case series. Such variability limits the strength of pooled estimates.

Second, follow-up durations differed considerably making comparison difficult and resulting in the transient early improvements in some severe-AD cohorts.

Third, stimulation parameters were differed among studies. Voltage varied widely (1–7 V), pulse width ranged from 60 to 210 µs, and frequency differed from 40 Hz to 130 Hz. Moreover, half of the studies (5/10) did not report programming duration at all, which restricts dose-response relationships.

Fourth, outcome measures were varied in reports. Only four studies provided sufficient quantitative data for ADAS-Cog meta-analysis, and motor/ADL outcomes were sparse and inconsistent, often limited to FIM or Barthel scores without standardized motor testing.

Finally, although the funnel plot for ADAS-Cog showed approximate symmetry, the small number of studies reduces the statistical power to detect publication bias or confirm true absence of heterogeneity.

### Future direction

future direction can be set in the below points:

`o`  Precision targeting using tractography to engage specific fornical sub-bundles.
`o`  Adaptive or closed-loop DBS that adjusts stimulation based on real-time physiological markers.
`o`  Age-stratified stimulation paradigms, addressing the opposing responses seen in older vs younger patients.
`o`  Stage-specific protocols, recognizing that severe AD may respond differently from mild AD.
`o`  Multi-target stimulation, such as combined fornix with nucleus basalis of Meynert DBS, which showed encouraging early results in severe AD (Xu, 2024).
`o`  Longer follow-up with harmonized parameters, to evaluate both early and late phases of network adaptation.

## Conclusion

while fornix DBS affects limbic and memory-related networks, it does not result in sustained cognitive or motor improvement. Meta-analytic findings showed pooled ADAS-Cog and MMSE effects centered around zero, with no influence from study design, age, or outlier effects based on subgroup, sensitivity, or meta-regression analyses. Short-term benefits occurred primarily in severe AD cohorts, whereas mild-AD trials showed no measurable response. Considerable heterogeneity in stimulation parameters and programming further underscores the need for standardized methodological frameworks. Overall, current evidence indicates that DBS-f produces biological activation without durable clinical benefit. Future progress will depend on optimized stimulation protocols, biomarker-driven patient selection, and emerging multi-target or adaptive neuromodulation strategies.

**Supplementary files:**

**Supplementary files:**

**Sup_1: Search_startegies**

**Sup_2: Full text exclusion and inclusion sheets Sup_3: risk of bias assessment sheets**

## Data Availability

All data produced in the present work are contained in the manuscript

