## Supplementary material for "Deep Brain Stimulation of the Fornix for Alzheimer’s Disease: A Systematic Review and Meta-Analysis of Cognitive and Motor Outcomes": Sup_3

Supp1: search strategies

Pubmed

(Alzheimer* OR "Alzheimer disease" OR dementia OR "cognitive decline" OR "mild cognitive impairment") AND ("deep brain stimulation" OR DBS OR "brain stimulation" OR "neuromodulation") AND (fornix OR "fornical region" OR "fornical area") AND (cognit* OR memory OR "executive function" OR "neuropsychological test*" OR "neurocognitive" OR "cognitive outcome*" OR "cognitive function*")

WOS:

TS=(Alzheimer* OR "Alzheimer disease" OR dementia OR "cognitive decline" OR "mild cognitive impairment")

AND

TS=("deep brain stimulation" OR DBS OR "brain stimulation" OR neuromodulation)

AND

TS=(fornix OR "fornical region" OR "fornical area")

AND

TS=(cognit* OR memory OR "executive function" OR "neuropsychological test" OR "neuropsychological tests" OR neurocognitive OR "cognitive outcome" OR "cognitive outcomes" OR "cognitive function" OR "cognitive functions")

Embase

('alzheimer*':ti,ab OR 'alzheimer disease':ti,ab OR dementia:ti,ab OR 'cognitive decline':ti,ab OR 'mild cognitive impairment':ti,ab) AND ('deep brain stimulation':ti,ab OR dbs:ti,ab OR 'brain stimulation':ti,ab OR neuromodulation:ti,ab) AND (fornix:ti,ab OR 'fornical region':ti,ab OR 'fornical area':ti,ab) AND (cognit*:ti,ab OR memory:ti,ab OR 'executive function':ti,ab OR 'neuropsychological test':ti,ab OR 'neuropsychological tests':ti,ab OR neurocognitive:ti,ab OR 'cognitive outcome':ti,ab OR 'cognitive outcomes':ti,ab OR 'cognitive function':ti,ab OR 'cognitive functions':ti,ab)

Scopus

TITLE-ABS-KEY (Alzheimer* OR "Alzheimer disease" OR dementia OR "cognitive decline" OR "mild cognitive impairment") AND TITLE-ABS-KEY ("deep brain stimulation" OR DBS OR "brain stimulation" OR neuromodulation) AND TITLE-ABS-KEY (fornix OR "fornical region" OR "fornical area") AND TITLE-ABS-KEY (cognit* OR memory OR "executive function" OR "neuropsychological test" OR neurocognitive OR "cognitive outcome" OR "cognitive function")
